# Music-Synchronized Gamma Light Stimulation Strengthens Theta-Gamma Coupling in Older Adults with Cognitive Impairment: Results from a Randomized Feasibility Trial

**DOI:** 10.64898/2026.11.06.26367319

**Authors:** Benjamin M. Kubit, Arun Asthagiri, Kelsie L. Lopez, Jinyu Wang, Corinna Parrish, Bowen Trebour, Ziyan (Annie) Zhao, Damla Yesil, Amira Toivonen, Aaron Jacobowitz, Brynna Quigley, Eva Xiaotong Wu, Ji Chul Kim, Edward Large, Psyche Loui

## Abstract

Noninvasive clinical innovations for restoring gamma activity via sound and light stimulation have the potential to improve memory and cognitive abilities in Alzheimer’s disease. We assessed whether pairing music with theta- and gamma-band light changes memory-supporting neural dynamics, and whether those changes track cognition. In an ongoing randomized feasibility trial, 38 older adults with cognitive impairment completed eight weeks of an audiovisual intervention. The treatment group (n = 19) listened to self-selected music while viewing light stimulation that consisted of a 39-Hz gamma-band signal modulated by delta- and theta-band musical rhythms. The control group (n = 19) listened to podcasts while viewing synchronized lights without the gamma frequency (delta-band only). Before and after the 8-week intervention, participants completed a face-name association task and the Alzheimer’s Disease Assessment Scale-Cognitive (ADAS-Cog). Individuals in the treatment group showed significant improvements on a face-name association task and the ADAS-Cog. Cluster-based permutation testing showed a larger increase in phase-amplitude coupling between acoustic theta phase and frontal EEG gamma amplitude in the treatment group compared to the control group. Across participants, increases in stimulus-driven theta to EEG-gamma phase-amplitude coupling were associated with improvements in memory. These preliminary results suggest that theta-gamma stimulation delivered through music and lights strengthens cross-frequency coupling, and may correspondingly strengthen associative memory.

## Introduction

Over 7 million people aged 65 years and older live with Alzheimer’s disease (AD) in the United States, with potential for that number to increase to 13.8 million by 2060 (“2025 Alzheimer’s Disease Facts and Figures,” 2025). AD is associated with a number of adverse behavioral and psychological symptoms, including increased stress, agitation, depression, low motivation, and cognitive deficits, such as memory loss and cognitive impairment (Tenchov et al., 2024). Underlying neurobiological mechanisms of AD have begun to be uncovered to help explain these behavioral and psychological changes.

Specifically, gamma-band activity has been shown to be altered in individuals with AD (Traikapi & Konstantinou, 2021). Prior literature has indicated that these altered gamma oscillations precede the development of amyloid-β (Aβ) protein deposits, a key biomarker for AD (Iaccarino et al., 2016).

Advances in understanding altered gamma oscillations as a biomarker of AD have motivated noninvasive clinical innovations for restoring high-frequency activity with gamma stimulation via targeted sounds and lights. In animal models of AD, auditory (Martorell et al., 2019), light (Adaikkan et al., 2019; Singer et al., 2018), and combined audiovisual (Martorell et al., 2019) gamma stimulation (i.e., lights and sounds at 40 Hz) were found to restore gamma-band neural activity in primary sensory brain areas and the hippocampus. In turn, restoration of gamma oscillations decreases Aβ-plaque deposits (Iaccarino et al., 2016) via astrocytic clearance within the glymphatic system (Murdock et al., 2024), and improves memory (e.g., spatial memory; Adaikkan et al., 2019; Martorell et al., 2019). In humans, pilot results from a 3-month intervention leveraging audiovisual stimulation of gamma demonstrated slowed hippocampal atrophy and improvements in memory performance of patients with mild AD following the intervention (Chan et al., 2022). Preliminary results from this pilot study suggest that targeting disrupted gamma neural activity with gamma sensory stimulation has potential to improve cognitive deficits in AD.

In addition to direct stimulation of gamma oscillations, targeting cross-frequency coupling between theta and gamma rhythms may be particularly effective at mitigating cognitive decline in aging, given the functional role of theta-gamma cross-frequency dynamics in working memory, item-sequence binding, and sequence learning (Canolty & Knight, 2010; Heusser et al., 2016; Lisman & Jensen, 2013; Tort et al., 2009). Theta-gamma phase-amplitude coupling (PAC) decreases in individuals with AD and mild cognitive impairment with respect to healthy controls and predicts deficits in working memory performance (Goodman et al., 2018). Prior work shows that noninvasive brain stimulation via transcranial Alternating Current Stimulation (tACS) can strengthen theta-gamma coupling and improve cognition in older adults (Alekseichuk et al., 2016; Reinhart & Nguyen, 2019). In adults aged 60 to 76, 25 minutes of tACS tuned to each participant’s own theta frequency increased working memory accuracy to the level of adults in their twenties (Reinhart & Nguyen, 2019). The benefit emerged during stimulation and lasted through the 50 minutes of testing that followed and was associated with restored theta-gamma PAC within the left temporal cortex and restored theta phase synchronization between temporal and prefrontal cortex.

Additionally, Grover et al. (2022) gave adults aged 65 to 88 four consecutive days of tACS during a word-recall task. Theta stimulation over the inferior parietal lobule improved working memory and gamma stimulation over the dorsolateral prefrontal cortex improved long-term memory. Each effect was still present one month later. Thus, selective changes to memory processes are possible through entrainment of theta and gamma brain rhythms.

Despite this promise, there are a number of important challenges in developing noninvasive methods for human interventions that potentially limit effectiveness, compliance, and scalability. One study in mice reported that 40 Hz light flicker did not engage native gamma oscillations in visual cortex, entorhinal cortex, or hippocampus, and that neither Aβ nor microglial morphology changed (Soula et al., 2023), a conclusion that other groups have disputed and that remains unsettled. Two further observations in that study bear on stimulus design. Hippocampal spike responses were weak, which suggests that light alone does not reach deep structures. And the mice avoided the 40 Hz flicker when given the choice between it and continuous light of the same intensity, with the avoidance accompanied by elevated hippocampal cholinergic activity. Auditory gamma carries its own constraint: amplitude modulation in the 30 to 40 Hz range is acoustically rough, so delivering gamma as sound alters the stimulus in ways listeners find unpleasant (Arnal et al., 2019). Response to auditory gamma stimulation also differs enough across humans that no single protocol suits everyone (Bolland et al., 2025), and methods such as tACS and transcranial Direct Current Stimulation (tDCS) require specialized equipment and trained administration, which limits at-home use and access for older adults.

By contrast, music is a widely accepted, low-cost, and scalable form of stimulation that effectively drives neural activity in multiple brain networks that are important for learning and memory (Chen et al., 2022; Harding et al., 2019; Henry et al., 2014; Loui et al., 2026). Music reliably drives brain activity in the delta (1-4 Hz) and theta (4-8 Hz) bands at frequencies associated with the beat level and the subdivisions of the beat in music, respectively (Ding et al., 2017; Doelling & Poeppel, 2015; Large et al., 2015). In addition, musical rhythms induce bursts of endogenous gamma (30-50 Hz) activity, suggesting that musical rhythms can be used to stimulate the gamma band (Fujioka et al., 2009; Snyder & Large, 2005). Neural synchronization to music therefore spans delta, theta, and gamma, the same bands disrupted in AD. Yet rhythmic synchronization to music is preserved in aging (Tichko, Page, et al., 2022), making it a candidate target for music-based intervention. Crucially, engaging with music is highly rewarding and pleasurable (Zatorre, 2015), and listening to familiar and enjoyable music drives brain activity in the dopaminergic reward system while increasing functional connections with the auditory system (Pereira et al., 2011; Salimpoor et al., 2013). In experience-sampling work, music accompanied a substantial share of everyday activities, and listeners reported more positive affect when the music was their own choice (Sloboda et al., 2001). Quinci and colleagues (Quinci et al., 2022) found that an eight-week music-based intervention engaged auditory and reward networks in older adults, suggesting that a sustained, music-based protocol in similar populations engages these systems. Leveraging the rewarding nature of music thus has the potential to increase compliance in AD interventions.

Music-based interventions (MBIs) address some of the symptoms associated with AD, including agitation and depression (Hillebrand et al., 2023; Van Der Steen et al., 2025). Whether music can also address the declines in memory and cognition associated with mild AD and mild cognitive impairment (MCI) is less clear, and whether music can do so through rhythmic synchronization of neural oscillations has not been directly tested. In the present two-arm randomized feasibility trial, participants completed an 8-week audiovisual intervention that is designed to rehabilitate neural oscillations using a combination of musical rhythms and synchronized light pulses (see Figure 1A for example of device used for light stimulation). The treatment group listened to self-selected music while receiving music-synchronized light stimulation in the delta (1-4 Hz) and theta (4-8 Hz) frequency ranges, along with amplitude-modulated bursts of gamma (39 Hz) intended to synchronize cross-frequency dynamics across those frequencies (hereafter referred to as delta-theta-gamma-driven light stimulation; Figure 1B and 1C). The control group listened to self-selected health education podcasts while receiving speech-synchronized light stimulation in the delta range alone (hereafter referred to as delta-driven light stimulation) (see Figure 1D for design of randomized feasibility trial). Because both arms received audio paired with synchronized light, the contrast isolates self-selected music combined with light targeting theta-gamma coupling. We hypothesized that this combination would strengthen stimulus-driven theta-gamma phase-amplitude coupling (PAC), and that the size of the change would correspond to improvement in cognition. We operationalized stimulus-driven theta-gamma PAC (hereafter referred to as theta-gamma PAC) by measuring PAC between the audio-derived theta signal and the EEG-derived gamma signal. Here we report feasibility, cognitive outcomes, and changes in theta-gamma PAC in an initial subset of participants from this ongoing randomized feasibility trial.

**Figure 1.**
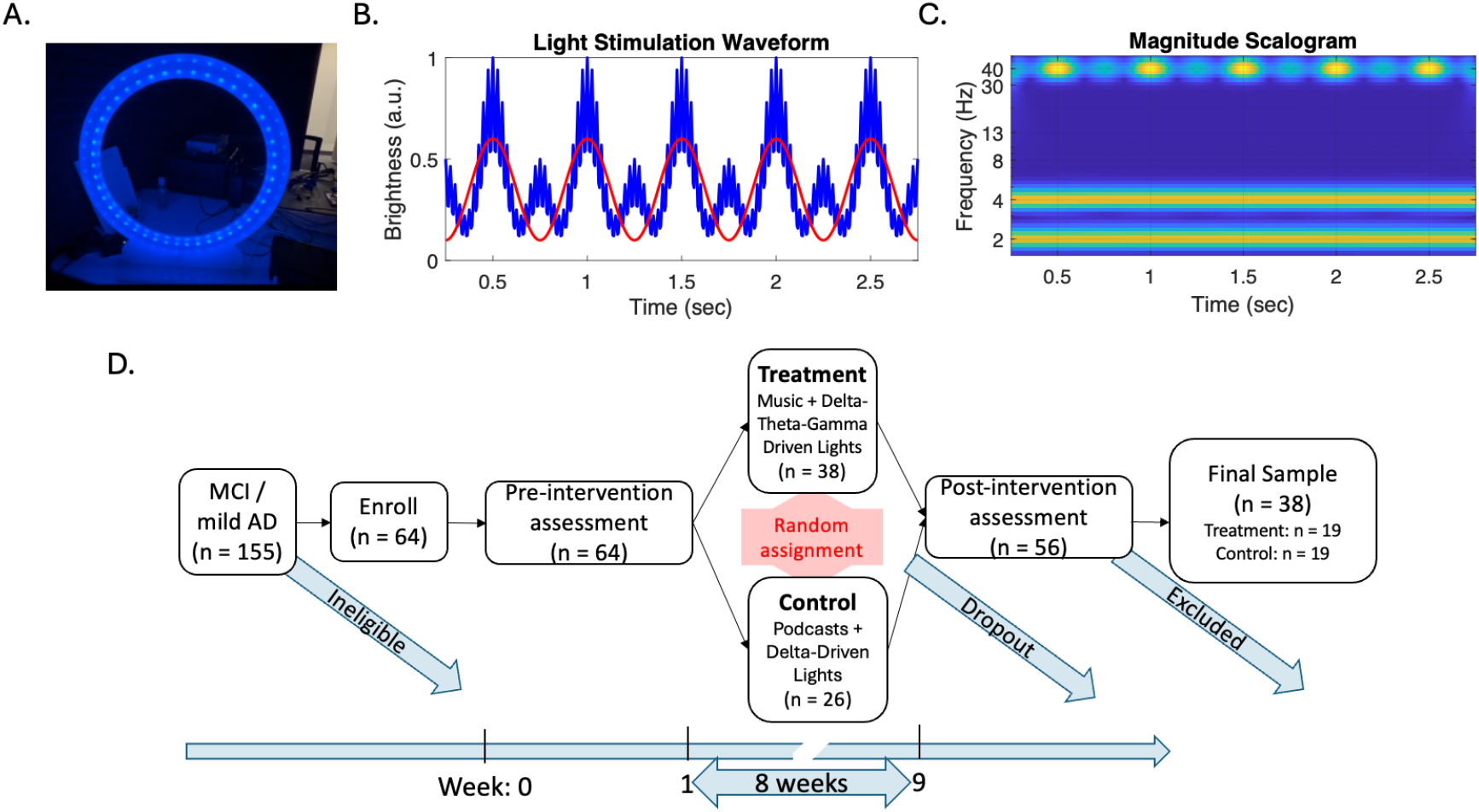
(A) Light stimulation device used in intervention. (B) The waveform and (C) time-frequency analysis of the light stimulation produced by the delta-theta-gamma-driven light for a 2-Hz rhythm. The red curve in B shows the delta wave (2 Hz) only. (D) Protocol outlining the design of a randomized feasibility trial from screening through to intervention completion.

## Results

### Compliance

On average, participants returned 9 days after finishing the 8-week intervention (range 1 to 39 days) and a total of 8 participants dropped out, 5 from the treatment group and 3 from the control group. The protocol asked for 30 minutes a day across 56 days, or 1,680 minutes in total. Both groups listened on a similar number of days (treatment M = 51.2, SD = 19.6; control M = 46.0, SD = 18.8; difference 5.2 days, 95% CI [-7.5, 17.8]; t(36) = 0.83, p = .413, d = 0.27), but treatment sessions lasted longer, averaging 46.0 minutes against 30.6 in the control group (treatment M = 46.0, SD = 22.2, control M = 30.6, SD = 4.8; difference 15.4 minutes, 95% CI [4.8, 26.0]; t(35) = 3.0, p = .006, d = 0.97). Control sessions clustered tightly around the 30-minute minimum, while treatment sessions ranged from 29.8 to 123.1 minutes. Participants in the treatment group exceeded 30 minutes of listening on more days than participants in the control group (treatment M = 49.4, SD = 19.3; control M = 29.9, SD = 17.1; difference 19.4 days, 95% CI [7.4, 31.4]; t(36) = 3.3, p = .002, d = 1.06; Figure S1A). The treatment group also listened for longer in total across the eight weeks (treatment M = 2,342, SD = 1,264; control M = 1,390, SD = 610; difference 952 minutes, 95% CI [300, 1,605]; t(36) = 3.0, p = .005, d = 0.96; Figure S1B). Eighteen participants logged listening on more than 56 days (11 in the treatment group and 7 in the control group).

Participants rated the lights positively in both groups, and adding theta and gamma components to the light did not make it less enjoyable. Ratings were slightly higher in the treatment group on both items, and the difference was not significant (enjoyable: treatment M = 6.9, SD = 1.8; control M = 5.9, SD = 2.6; difference 1.0, 95% CI [−0.6, 2.4]; *t*(34) = 1.21, *p* = .235, *d* = 0.40; stimulating: treatment M = 6.9, SD = 1.9; control M = 6.0, SD = 2.6; difference 0.9, 95% CI [−0.7, 2.4]; *t*(33) = 1.15, *p* = .259, *d* = 0.39).

### Stimulus Validation

The combination of music and light stimulation was effective at increasing gain in the delta and theta frequencies while listening to both “Under the Boardwalk” by The Drifters and “Graceland” by Paul Simon (collapsed across both light stimulation conditions; Figure 2A). Further, delta and theta frequencies in the acoustic and EEG signals synchronized, as shown through a consistent phase-locking during both songs (again, collapsed across light conditions; Figure 2B). Phase-locking peaked at the beat and subdivision frequencies of each song: 2.05 and 4.09 Hz for “Under the Boardwalk” and 1.96 and 3.92 Hz for “Graceland.”

**Figure 2.**
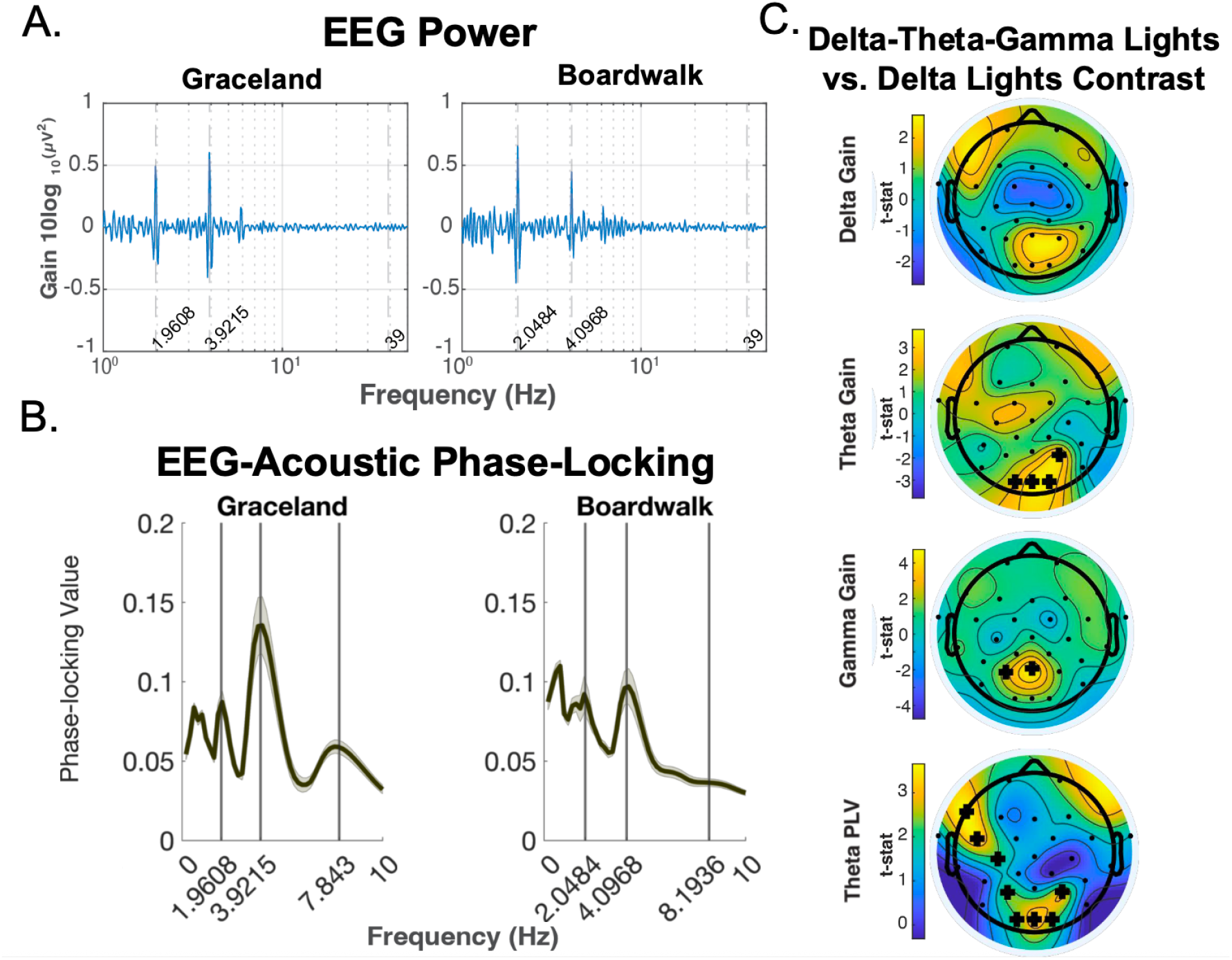
(A) EEG spectral gain and (B) phase-locking value (PLV) of delta and theta frequencies while listening to “Under the Boardwalk” and “Graceland” during light stimulation in the pre-intervention EEG session. (C) Topographic plots of gain and phase-locking contrasts between delta-theta-gamma-driven light stimulation and delta-driven light stimulation conditions in the pre-intervention EEG session.

Contrasts of delta-theta-gamma-driven light stimulation and delta-driven light stimulation from the pre-intervention EEG task revealed more theta brain oscillations in parietal and occipital regions and more gamma brain oscillations in parietal regions, while viewing the delta-theta-gamma-driven light stimulation compared to the delta-driven light stimulation. A cluster-based permutation test across electrodes showed higher theta gain over electrodes P4, O1, Oz, and O2 (cluster-corrected p = .017; within-cluster d_z_ = 0.65, 95% CI [0.32, 1.09]), and higher gamma gain over electrodes P3 and Pz (cluster-corrected p = .035; within-cluster d_z_ = 0.72, 95% CI [0.44, 1.08]) when viewing the delta-theta-gamma-driven light stimulation compared to the delta-driven light stimulation. There was no difference in delta gain between the two conditions (cluster-corrected p = .098). Additionally, widespread increases in theta PLV were found for the delta-theta-gamma-driven light stimulation compared to the delta-driven light stimulation, as indicated by the following significant electrodes following a cluster-based permutation test: F7, FC5, C3, P3, P4, O1, Oz, and O2 (cluster-corrected p = .004; within-cluster d_z_ = 0.67, 95% CI [0.34, 1.09]; Figure 2C).

### Behavioral Assessments

An ANOVA on face-name scores (n = 37; one treatment-group participant was excluded for equipment failure during the task) revealed a significant main effect of Session (*F*(1,35) = 7.75, *p* = .009, ηp² = .18), with post-hoc analyses revealing a significant within-group change from pre- to post-intervention on the face-name association task for the treatment group (*t*(17) = 2.8, *p* = .013, *d*_z_ = 0.66; Figure 3A). The main effect of Intervention did not reach significance (F(1,35) = 2.44, p = .128, ηp² = .07), but there was a marginal Intervention x Session interaction on the face-name association task (*F*(1,35) = 3.60, *p* = .066, ηp² = .09), such that greater improvements in associative memory abilities were found in the treatment group following the 8-week intervention. The between-group comparison of change scores gave t(35) = 1.90, p = .066, d = 0.62, 95% CI [−0.04, 1.28]. Scores were higher on the face-name association task following the 8-week treatment group intervention (pre: *M* = 63.5, *SD* = 14.1; post: *M* = 73.4, *SD* = 13.1), indicating improvements in associative memory abilities. There was no significant within-group difference in scores on the face-name association task from pre- to post-intervention in the control group (*t*(18) = 0.8, *p* = .438, *d*_z_ = 0.18; pre: *M* = 74.1, *SD* = 15.1; post: *M* = 75.9, *SD* = 14.7).

**Figure 3.**
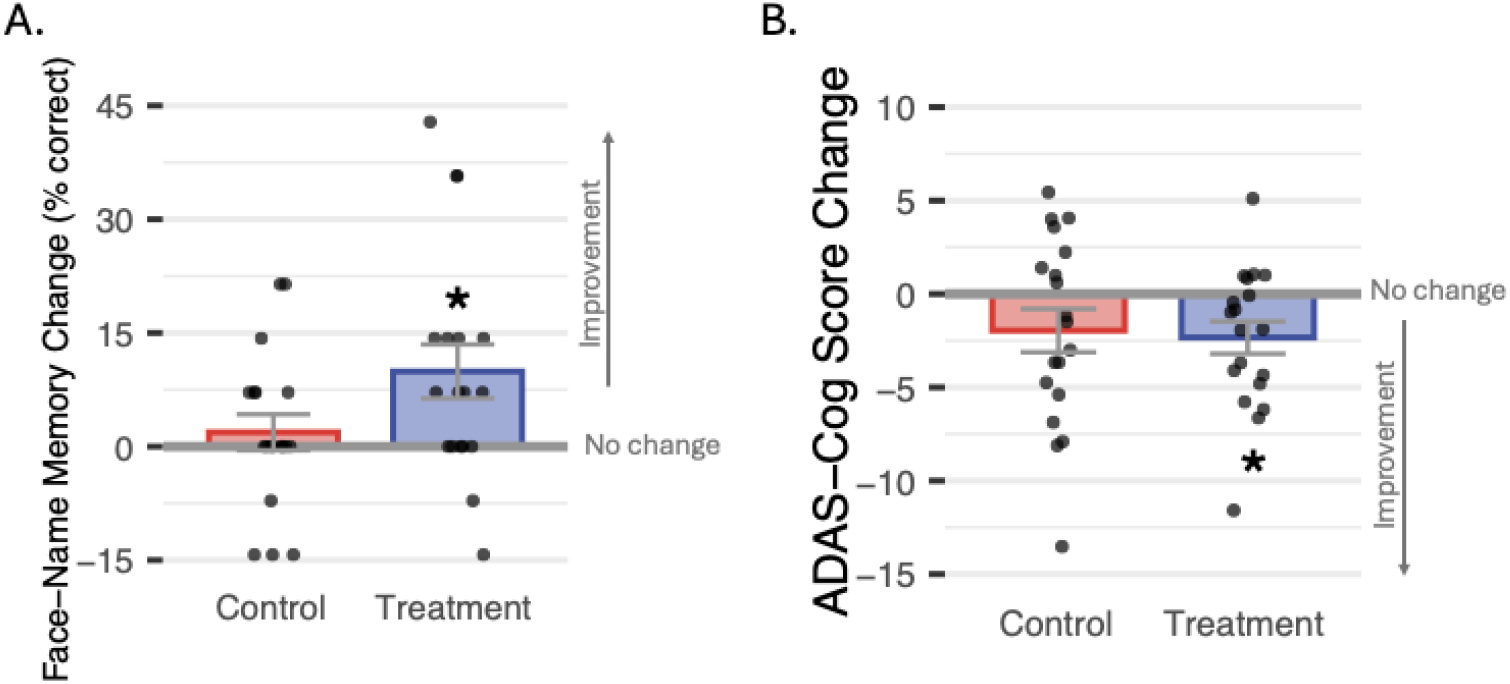
Change in scores on the (A) face-name association task and (B) ADAS-Cog after the 8-week intervention stratified by intervention protocol. Positive percent change indicates improvement on the face-name association task. Negative score change indicates improvement on the ADAS-Cog. Asterisks indicate significance of t-tests (* *p* < .05; ** *p* < .01; *** *p* < .001).

The same ANOVA model predicting ADAS-Cog scores, where lower scores indicate less impairment, showed a similar pattern (n = 38). There was a significant main effect of Session (*F*(1,36) = 8.79, *p* = .005, ηp² = .20), with post-hoc analyses revealing a significant within-group change from pre- to post-intervention on the ADAS-Cog for the treatment group (*t*(18) = -2.7, *p* = .015, *d*_z_ = 0.62; Figure 3B). The main effect of Intervention did not reach significance (F(1,36) = 0.93, p = .342, ηp² = .03) but, unlike face-name results, there was no Intervention x Session effect on ADAS-Cog scores (*F*(1,36) = 0.07, *p* = .796, ηp² = .002). Scores were lower on the ADAS-Cog following the 8-week treatment intervention (pre: *M* = 15.1, *SD* = 9.1; post: *M* = 12.8, *SD* = 7.5), indicating improvements in cognition. There was no significant within-group difference in scores on the ADAS-Cog from pre- to post-intervention in the control group (*t*(18) = -1.7, *p* = .108, *d*_z_ = 0.39; pre: *M* = 12.7, *SD* = 6.2; post: *M* = 10.8, *SD* = 6.9). Pre- and post-intervention descriptive statistics on the additional set of neuropsychological tests are presented in Table S1.

### Theta-Gamma Phase-Amplitude Coupling

Our audiovisual stimulus carried theta-gamma coupling by design (see Figure 1). We used EEG to ask whether our stimulation altered brain dynamics at the hypothesized frequencies, with theta phase determined from the audio and gamma amplitude driven by the light, over the course of the intervention. Thus, audio-derived theta to EEG-derived gamma phase-amplitude coupling (referred to as theta-gamma PAC) served as the primary EEG outcome measure for the intervention. We measured theta-gamma PAC separately for each electrode in the pre- and post-intervention EEG testing sessions. Theta-gamma PAC changed in opposite directions in the two groups (Figure 4A; EEG data were available for 34 out of the 38 participants). In the treatment group (n = 17), PAC increased from pre- to post-intervention over frontal electrodes. In the control group, PAC decreased across most of the scalp. We compared the two changes using a cluster-based permutation test across electrodes, which revealed a significantly greater change in PAC for the treatment group over a frontal cluster (Fp1, Fp2, F7, F3; cluster-corrected p = .014; within-cluster d = 1.05, 95% CI [0.31, 1.68]; Figure 4B). The strengthened coupling in the treatment group is visible in the distribution of gamma amplitude over theta phase, shown for “Graceland” in Figure 4C: gamma amplitude became more concentrated at particular theta phases over the course of the intervention, and the same concentration did not develop in the control group (n = 17). Delta and theta gain and theta phase-locking showed the same Intervention x Session pattern over central-parietal and occipital clusters; gamma gain did not differ between groups (Figure S2).

**Figure 4.**
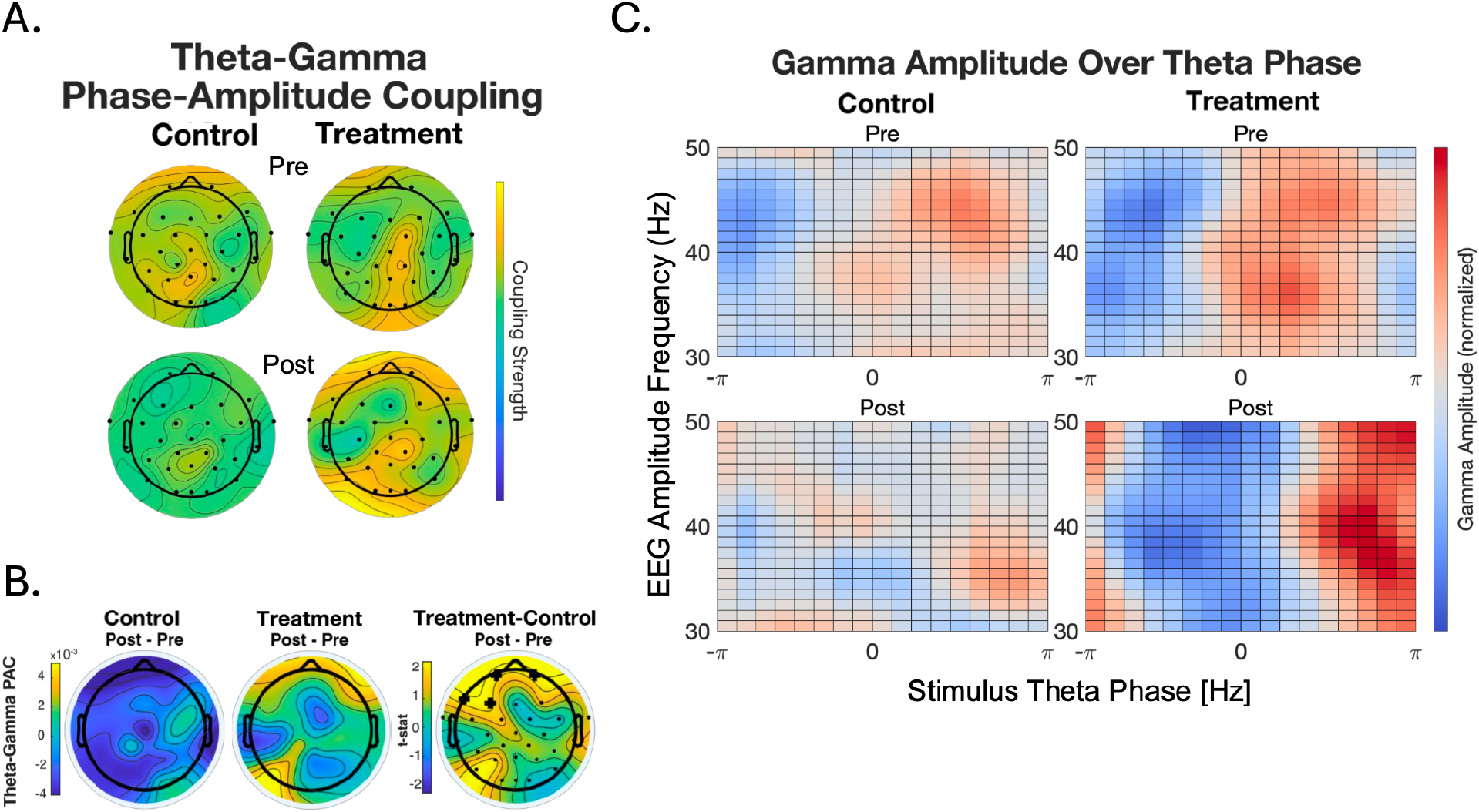
(A) Topographic plots of stimulus driven theta to EEG-gamma phase-amplitude coupling strength during the pre-intervention and post-intervention sessions for control and treatment groups. (B) Topographic plots of change in theta-gamma phase-amplitude coupling strength from pre-intervention to post-intervention shown for the control and treatment groups and a contrast between the control and treatment groups. Significant electrodes in the contrast plot determined from a cluster-based permutation test are marked with “+”. (C) Gamma amplitude over theta phase while listening to the song “Graceland” during the pre-intervention and post-intervention EEG session for the control and treatment groups.

### Association Between Coupling and Memory

Behavioral performance and EEG data were available for 33 out of the 38 participants. Across both intervention groups, change in stimulus-driven theta-gamma PAC from pre- to post-intervention was associated with change in face-name association score (Pearson r = 0.39, permutation p = .028, 95% CI [0.10, 0.63]; Figure 5A). Within groups, the association reached significance in the control group (Pearson r = 0.63, permutation p = .008, 95% CI [0.27, 0.86]) but not in the treatment group (Pearson r = 0.21, permutation p = .430, 95% CI [−0.25, 0.56]), and the slopes were not significantly different (interaction F(1,29) = 3.57, p = .069, ηp² = .11). Change in PAC was not associated with change in ADAS-Cog score (Pearson r = −.05, permutation p = .776, 95% CI [−0.36, 0.22]; Figure 5B).

**Figure 5.**
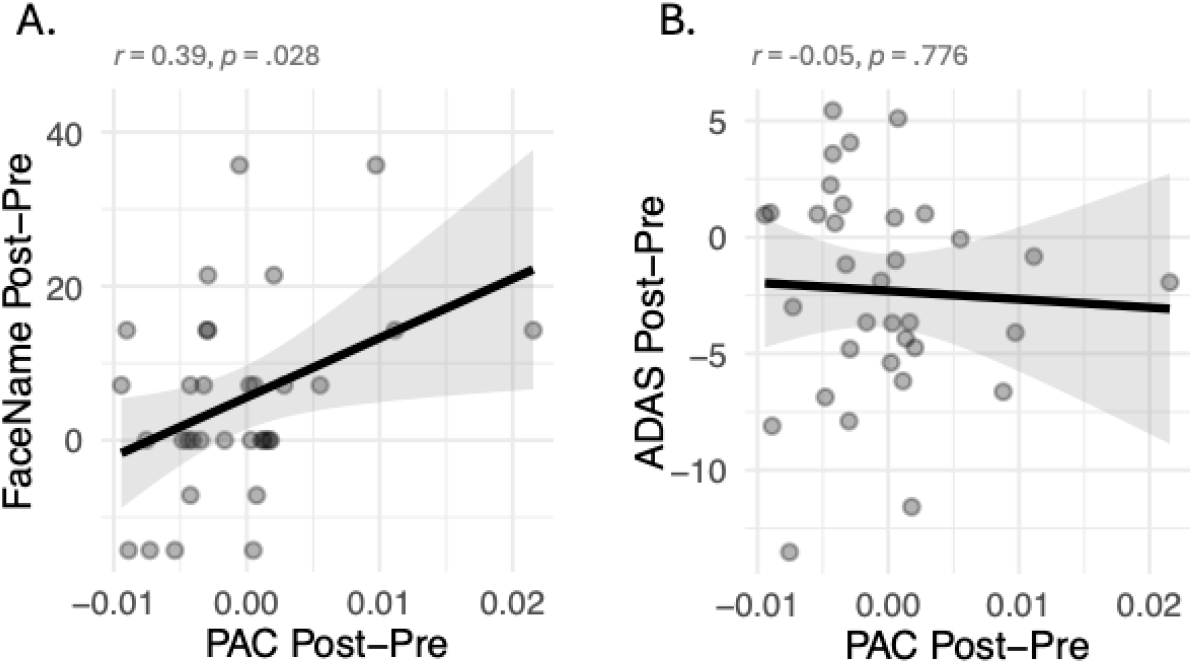
Associations between change in theta-gamma phase-amplitude coupling and change in scores on the (A) face-name association task and (B) ADAS-Cog from pre-intervention to post-intervention.

## Discussion

In this preliminary report we present interim results from an 8-week music-based intervention combining self-selected music with synchronized light, in adults with MCI and mild AD. Five findings emerged. Participants completed both protocols at high rates, and the treatment group listened for longer than the control group. Both groups rated the lights positively throughout. Face-name and ADAS-Cog scores improved across the sample, with the within-group improvement reaching significance only in the treatment group. Stimulus-driven theta to EEG-gamma phase-amplitude coupling (theta-gamma PAC) diverged between the groups over frontal electrodes by the end of the 8 weeks, decreasing in the control group and holding steady or increasing in the treatment group. The magnitude of that change over the course of the intervention positively correlated with improvements in associative memory.

Compliance was high across both the treatment and control groups. The treatment group listened and watched for, on average, 952 more minutes across the eight weeks than the control group. The difference was not in how often people used the device, since both groups listened and watched on a similar number of days (treatment for 51 days and control for 46, on average), but in how long each session lasted: 46 minutes on average in the treatment group compared to 31 minutes in the control group.

Participants in the treatment group consistently chose to use the intervention more than protocol required, suggesting that listening to music was more engaging than health education podcasts. Importantly, adding theta and gamma components to the light did not make it less enjoyable. While discomfort with gamma-augmented lights and sounds has limited other interventions, in the present treatment protocol, participants kept rating the lights positively after two months of daily use. The two protocols differ in both their audio and their light, so we cannot attribute the results to music alone, though music is rewarding in ways health education podcasts are not (Zatorre, 2015), and people choose to listen in daily life, particularly when the music is their own selection (Sloboda et al., 2001). As a result, interventions that feature music may be better at promoting compliance within clinical populations who may suffer from low motivation.

The audiovisual stimulation worked as designed. In the pre-intervention session, the delta-theta-gamma-driven light produced higher gain at the theta and gamma frequencies than the delta-driven light, over parietal and occipital electrodes, and more widespread phase-locking to the theta frequency. As expected, gain in the delta frequency did not differ between the conditions since both included the delta visual component. The added theta and gamma components accomplished what they were designed to do.

Both ADAS-Cog and face-name memory scores improved across the sample over the eight weeks, and neither showed a significant Intervention x Session interaction. However, the within-group improvements reached significance only in the treatment group, suggesting potential improvements in cognition and associative memory abilities. The treatment group’s face-name association memory improved 9.9 percentage points (p = .013, *d*_z_ = 0.66) versus 1.8 in controls (p = .438), a medium-sized effect that was not significantly different between groups at our interim sample size (p = .066, d = 0.62). A recent review showed that nonpharmacological interventions can be effective in improving ADAS-Cog, particularly repetitive transcranial magnetic stimulation (rTMS), although the authors note that research using this outcome measure is still limited (Cui et al., 2026). Much of the literature that notes improvements in face-name association abilities in AD relies on interventions that directly train cognition and memory during the intervention itself (Bier et al., 2008; Clare et al., 2002). Here, face-name scores improved in the treatment group without any direct training of cognition or memory, and we observed a parallel change in cross-frequency coupling.

Stimulus-driven theta-gamma PAC was successfully modulated by the intervention in the treatment group but not the control group in a frontal cluster (Figure 4A). The two groups moved in opposite directions. Coupling decreased in 14 of 17 control participants, across most of the scalp, and increased over the frontal electrodes in 13 of 17 treatment participants (Figure 4B). Reduced theta-gamma coupling in the brain is a feature of MCI and AD (Goodman et al., 2018), and the control group’s decline moves in that direction. Whether the difference in coupling reflects disease-related change over 8 weeks or an effect of the delta-driven control protocol itself cannot be separated with this design.

The pattern of results suggests the intervention may have preserved, rather than enhanced, coupling.

Theta-gamma coupling is thought to serve the binding of items in memory, which is what face-name recognition requires, while the ADAS-Cog aggregates orientation, language, praxis, and word recall. The difference between tasks is reflected in our results. The interaction effect size was larger for face-name association (ηp² = .09 against .002), and change in coupling corresponded with change in face-name (r = 0.39, p = .028) score but not with change in ADAS-Cog (r = -0.05). Within groups, the association reached significance in the control (*r* = 0.63, p = .008) but not the treatment group (*r* = 0.21), though the slopes did not reliably differ (interaction *p* = .069). Our finding therefore describes a relationship between coupling and memory in this population; further testing is needed to identify the specificity of this mechanism.

### Limitations and Future Directions

This is an interim analysis of an ongoing Stage I trial per the NIH Stage Model. Results reported here may change once the full sample is collected. In the current sample, the treatment group was more impaired than the control group at baseline on both behavioral measures, scoring 10.6 percentage points lower on the face-name association task and 2.4 points higher on the ADAS-Cog, and also scored lower on the TICS. Change scores are difficult to interpret against a baseline difference of this size, since the group starting further from ceiling has more room to improve. The group by session interactions did not reach significance on either behavioral measure. Randomization should balance the arms as enrollment continues, and the full sample will determine whether the difference between groups holds.

We measured coupling between the theta phase of the stimulus and the gamma amplitude of the EEG signal. Our approach differs from the coupling between theta and gamma within the brain that is reduced in AD and MCI (Goodman et al., 2018) and that tACS can restore (Reinhart & Nguyen, 2019). Whether the two are related is an open question. A brain that more faithfully reproduces an externally imposed coupling may also couple its own rhythms more strongly. One reason the two measures might diverge is that our theta frequency came from the music rather than from the participant. The beat subdivisions of the two songs fell at 4.09 and 3.92 Hz for every participant, whereas Reinhart and Nguyen (2019) tuned stimulation to each participant’s own theta frequency. If endogenous theta varies across people, a fixed stimulus theta will sit closer to the preferred theta for some participants than others, which could weaken the link between what the stimulus imposes and what the brain does on its own. Previous work has reported similar individual variability in response to gamma stimulation (Bolland et al., 2025).

In the present design, the treatment protocol paired music with theta-gamma stimulation, whereas the control protocol paired speech with delta-rate stimulation. Future work should independently cross auditory carrier (music vs. speech) with visual stimulation pattern (delta-theta-gamma-driven lights vs. delta-driven lights) to test for contributions of both as separate, but possibly synergistic effects on associative memory in cognitive impairment.

## Conclusion

Eight weeks of an MBI paired with delta, theta, and gamma light stimulation strengthened stimulus-driven theta-gamma PAC over frontal electrodes, while coupling declined over the same period in an active control. Associative memory and ADAS-Cog scores improved within the treatment group, and the change in coupling tracked the change in associative memory. We interpret these results as support for theta-gamma coupling as a candidate mechanism for the effect of rhythmic audiovisual stimulation on memory in MCI and mild AD.

## Methods

### Participants

Participants, recruitment, and eligibility criteria are those reported in Kubit et al. (2025); the full criteria are registered at ClinicalTrials.gov (NCT05984524). We recruited adults aged 50 to 90 from the metropolitan Boston area through paid advertisements and community outreach events. The protocol was approved by Northeastern University’s Institutional Review Board 19-03-20. A total of 155 individuals were assessed for eligibility. Those who indicated interest were screened by telephone interview with the Clinical Dementia Rating scale (CDR; Morris, 1997), and those with a CDR score of 0.5 to 2 were screened further. We also required that participants be willing and able to: 1) consent to participation, assessed with the UCSD Brief Assessment of Capacity to Consent (UBACC; Jeste et al., 2007); 2) complete the intervention and all assessments across the 8-week protocol; and 3) avoid new treatments other than the assigned intervention. We excluded individuals for: 1) changing dosage of cholinesterase inhibitors or psychotropic medication within 6 weeks of screening; 2) history of psychotic or schizophrenic episodes, major neurologic diagnosis (Parkinson’s, stroke, brain injury, epilepsy), or other conditions that might impair cognition or confound assessments; 3) history of chemotherapy treatment within the past 10 years; 4) recent (within the last 3 months) serious physical trauma or diagnosis of serious chronic health condition requiring medical treatment and monitoring; and 5) having participated in another intervention study within the past 30 days. Seven participants did not report normal hearing: four used hearing aids, one reported slight loss in one ear, and two described age-related loss. At the pre-intervention session we confirmed that every participant could hear the music and podcasts clearly through headphones and could follow normal conversation. Participants who used hearing aids removed them for the EEG sessions and confirmed they could still hear the stimuli through the headphones.

Of the 155 assessed, 91 were excluded, and the remaining 64 were randomized into the two intervention protocols (treatment and control). Eight participants dropped out during the 8-week intervention, leaving 56 who completed all aspects of it. Eighteen of those 56 were excluded from the present analyses. Thirteen did not receive the intervention in the form described under Materials: seven completed it before the final light stimulus was delivered by the manufacturer, and six completed it before subsequent corrections to the light stimulus, with their assessment data collected outside the laboratory.

Two were enrolled through a scoring error on the prescreen and did not meet criteria for MCI. The remaining three listened on too few days to meet the compliance threshold. Figure 6 gives the full flow of participant enrollment and study completion, including further information on ineligibility and exclusion. Thirty-eight older adults were included in the present analyses (see Table 1 for demographic information), but data collection is ongoing. Nineteen participants completed the treatment protocol and 19 completed the control protocol. EEG data were collected for 34 participants and 33 out of 38 participants had both behavioral performance and EEG.

**Figure 6.**
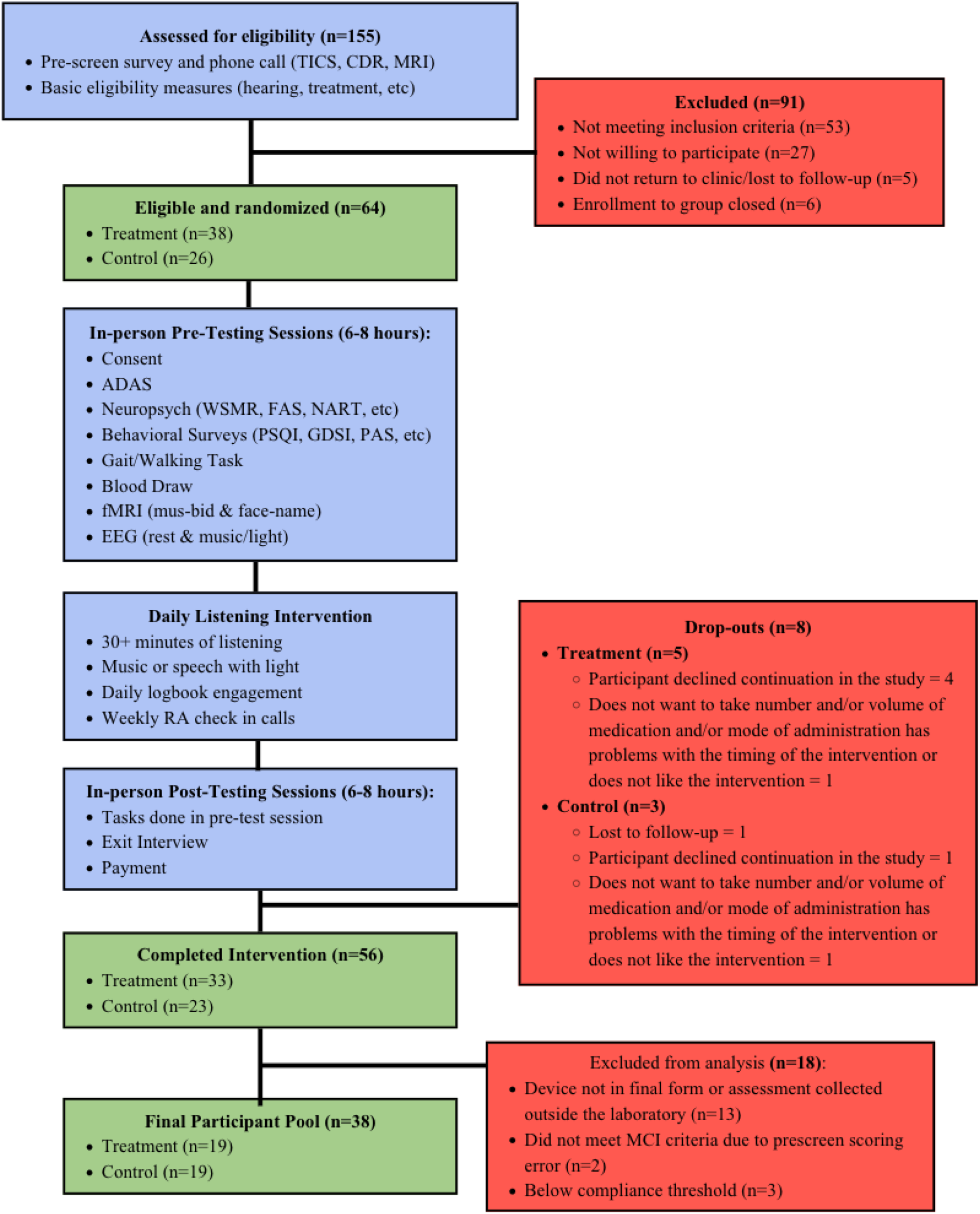
Flow chart of randomized feasibility trial enrollment, protocol, and study completion metrics.

**Table 1.**
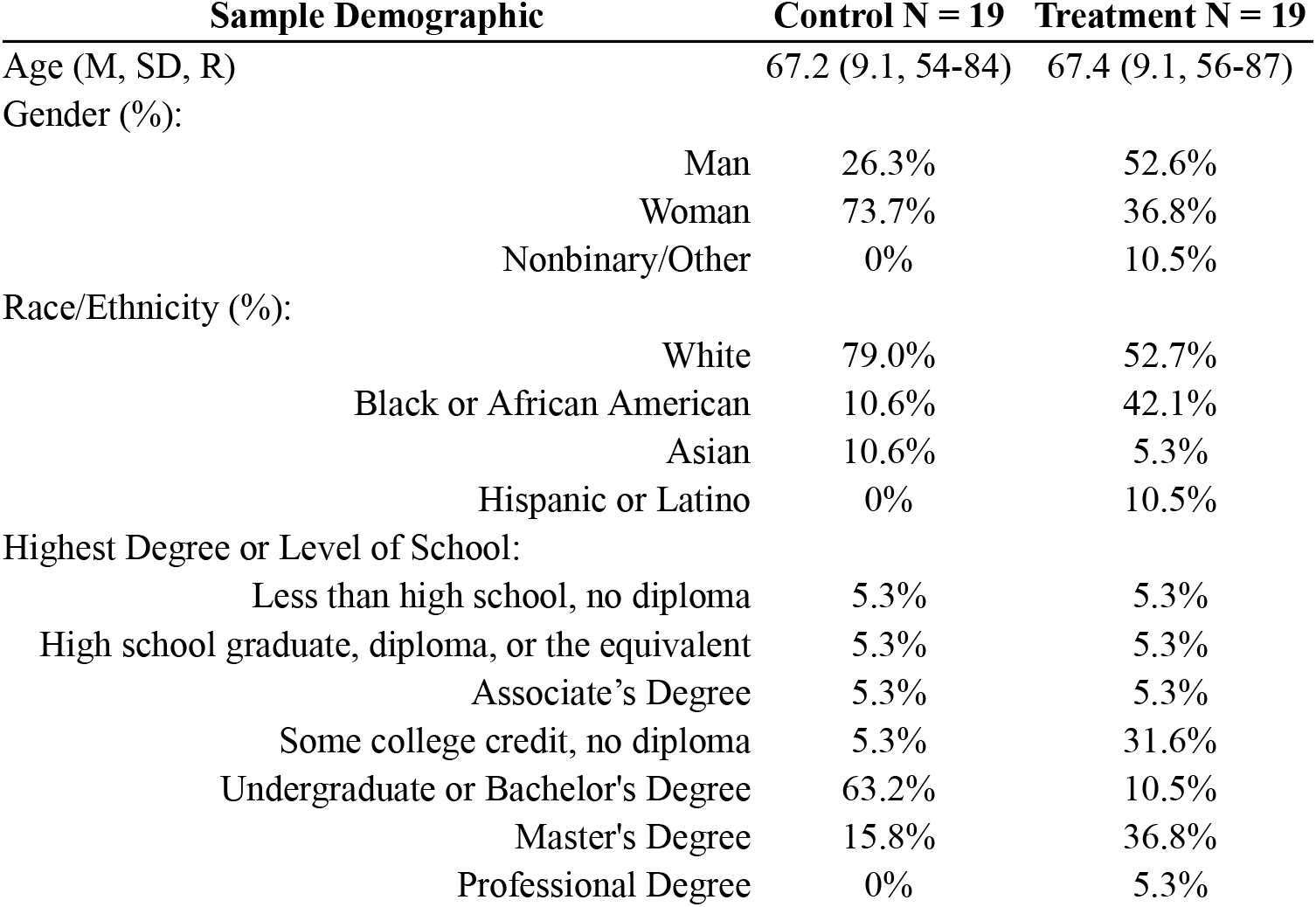
Control and Treatment groups sample demographics. M = Mean; SD = Standard Deviation; R = Range; % = percent of group.

| **Sample Demographic** | **Control N = 19** | **Treatment N = 19** |
|---|---|---|
| Age (M, SD, R) | 67.2 (9.1, 54-84) | 67.4 (9.1, 56-87) |
| Gender (%): |  |  |
| Man | 26.3% | 52.6% |
| Woman | 73.7% | 36.8% |
| Nonbinary/Other | 0% | 10.5% |
| Race/Ethnicity (%): |  |  |
| White | 79.0% | 52.7% |
| Black or African American | 10.6% | 42.1% |
| Asian | 10.6% | 5.3% |
| Hispanic or Latino | 0% | 10.5% |
| Highest Degree or Level of School: |  |  |
| Less than high school, no diploma | 5.3% | 5.3% |
| High school graduate, diploma, or the equivalent | 5.3% | 5.3% |
| Associate’s Degree | 5.3% | 5.3% |
| Some college credit, no diploma | 5.3% | 31.6% |
| Undergraduate or Bachelor's Degree | 63.2% | 10.5% |
| Master's Degree | 15.8% | 36.8% |
| Professional Degree | 0% | 5.3% |

Participants in the two groups were compared on a number of demographic factors relevant to the intervention (see Table 2). We found no significant group differences across measures of musical sophistication (GMSI-MS; Müllensiefen et al., 2014) and music reward sensitivity (eBMRQ; Cardona et al., 2022; Mas-Herrero et al., 2013), and performance on Clinical Dementia Rating (CDR; Morris, 1997) and Mini-Mental State Examination (MMSE; Folstein et al., 1975) was consistent across both groups. A significant difference was found for scores on the Telephone Interview for Cognitive Status (TICS; Brandt et al., 1988), such that the treatment group had lower scores than the control group on this

**Table 2.** Assessment of inter-group differences on key demographic factors. M = Mean; SD = Standard Deviation; % = percent of group. GMSI-MS = Goldsmiths Musical Sophistication Index - Musical Sophistication; eBMRQ = Extended Barcelona Music Reward Questionnaire; CDR = Clinical Dementia Rating scale; TICS = Telephone Interview for Cognitive Status; MMSE = Mini-Mental State Examination. Asterisks indicate significance of t-tests (* p < .05; ** p < .01; *** p < .001). CDR ranges from 0 to 3, where 0 = normal, 0.5 = very mild dementia, 1 = mild dementia, 2 = moderate dementia, 3 = severe dementia. TICS ranges from 0 to 41, where < or equal to 30 is cognitively impaired. MMSE ranges from 0 to 30, with a higher score indicating less impairment.

| Measure | Control | Treatment | Inter-group difference |
|---|---|---|---|
| Handedness: Right; n (%) | 19 (100%) | 14 (74%) | X<sup>2</sup>=3.7; p=.055 |
| Employment Status: Retired; n (%) | 11 (58%) | 8 (42%) | X<sup>2</sup>=0.4; p=.516 |
| Years Practicing Music: 0-2 Years; n (%) | 12 (63%) | 11 (58%) | X<sup>2</sup>=0.0; p=1.00 |
| Income: less than \$50,000; n (%) | 10 (53%) | 15 (79%) | X<sup>2</sup>=1.9; p=.171 |
| Neurological/Psychological Disorders: No; n (%) | 13 (68%) | 14 (74%) | X<sup>2</sup>=0.0; p=.000 |
| Normal Hearing: Yes; n (%) | 16 (84%) | 15 (79%) | X<sup>2</sup>=0.0; p=.000 |
| GMSI-MS: M (SD) | 65.8 (19.5) | 67.7 (19.2) | t=0.3; p=.764 |
| eBMRQ: M (SD) | 90.7 (15.4) | 94.6 (14.1) | t=0.8; p=.422 |
| CDR; n (%) 0.5 | 16 (84%) | 14 (74%) | X<sup>2</sup>=0.8; p=.670 |
| 1 | 2 (11%) | 4 (21%) |  |
| 2 | 1 ( 5%) | 1 ( 5%) |  |
| TICS; M (SD) | 34.2 (2.6) | 31.2 (3.4) | t=-3.0; p=.005** |
| MMSE; M (SD) | 27.1 (2.2) | 26.5 (2.7) | t=-0.8; p=.439 |

### Procedure

After determining eligibility, participants completed a pre-intervention assessment in the laboratory to establish a baseline on neuropsychological and neuroimaging measures. They were then randomly assigned to one of the two 8-week intervention protocols, and returned to the laboratory as soon as possible for a post-intervention assessment on the same measures. The assessments used are described below. Participants also completed structural and functional MRI at both sessions, and the face-name association task was administered in the scanner. We report the behavioral and electrophysiological outcomes here. The imaging analyses are ongoing and will be reported separately. The full battery administered at both sessions is listed in the trial registration (NCT05984524).

#### Pre-Intervention Assessment

Prior to random assignment to one of two intervention protocols, participants completed a series of pre-intervention assessments in the lab. The pre-intervention session took approximately 7 hours to complete and participants were compensated $20 per hour. The behavioral assessments and neuroimaging tasks completed by participants are outlined below. The additional collection of neuropsychological tests completed by participants is outlined in the supplemental materials.

##### Face-Name Association Task

We used the face-name association task (Sperling et al., 2001) to measure associative memory. Face-name associative memory performance on the task has been related to amyloid burden in elderly adults (Rentz et al., 2011) and associative recognition on this task is impaired in subjective cognitive decline and mild cognitive impairment (Polcher et al., 2017). The task also indexes the binding of item pairs, which is the process theta-gamma coupling is thought to support. Participants studied faces paired with names, and were then tested on each face against two names in a two-alternative forced choice.

Participants studied 30 face-name pairs, 16 female and 14 male, across 8 encoding blocks of 7 trials each. Four blocks presented 28 pairs a single time (novel condition), and the other four presented the remaining 2 pairs, one female and one male, 14 times each (repeated condition), which gives a within-subject contrast for encoding-related activity. Each face appeared for 4.75 s, followed by a 0.95 s fixation cross. Immediately after the final encoding block, participants were tested on 14 of the studied faces, 12 novel and 2 repeated. Each face appeared with the name it had been studied with and with one foil. Participants had 6.65 s to respond with a button press. Scores are percent correct and range from 0 to 100 (chance performance at 50). Data from one treatment-group participant were lost to equipment failure, leaving 37 participants for the face-name analyses.

##### Alzheimer’s Disease Assessment Scale - Cognitive (ADAS-Cog)

We used the 14-item Alzheimer’s Disease Assessment Scale, Cognitive (ADAS-Cog; (Mohs et al., 1997; Rosen et al., 1984)) to measure cognitive decline. The test takes approximately 30 minutes to complete in the laboratory. The research team was trained on the standardized administration procedure and met regularly to discuss unclear cases. The scale comprises the 11 items of the original version (Rosen et al., 1984) plus delayed word recall, number cancellation, and maze completion (Mohs et al., 1997). Scores range from 0 to 90, and higher scores indicate greater impairment.

The ADAS-Cog included a total of 14 cognitive tasks. The Word Recall task required the participant to recall a list of 10 words, and Delayed Word Recall required them to recall the same list at the end of administration. The Commands task required the participant to act out a series of commands from the experimenter (anywhere from one to five steps in the command). Constructional praxis involved replicating a geometric shape shown to the participant. The Naming Objects and Fingers task asked the participant to label a series of twelve objects and the fingers on their dominant hand. Ideational Praxis required the participant to go through the steps of sending a letter to themselves. The Orientation task asked the participant to indicate knowledge of facts such as the date, time, season, etc. Word Recognition involved recognizing a subset of previously seen words from a larger set of words. The Remembering Task Instructions assessment asked the experimenter to score the participant’s ability to remember the instructions of the Word Recognition task. The Spoken Language Ability task was a ten-minute open-ended conversation between the participant and the experimenter to assess spoken language ability. Relatedly, the Comprehension of Spoken Language assessed the language comprehension during this open-ended conversation. Simultaneously, the experimenter assessed Word Finding during the conversation, or difficulty in finding the words that they want. In the Maze task, participants completed a series of mazes. Finally, the Number Cancellation task asked participants to cross out target numbers within a large matrix of randomly arranged numbers.

##### Electroencephalography (EEG)

Participants listened to two songs, “Under the Boardwalk” (2 minutes 40.2 s; The Drifters, 1964) and “Graceland” (4 minutes 46.5 s; Paul Simon, 1986), each presented once with delta-theta-gamma-driven light stimulation and once with delta-driven light stimulation, for four listening trials per session. The light stimulation conditions are described under Materials. Audio was presented over headphones at a level that was comfortable to the participant. Trial order was counterbalanced across participants and across sessions.

Continuous EEG was recorded in an electrically shielded, sound-attenuated chamber at Northeastern University using a Brain Products actiCHamp system with PyCorder software and 32 gel-based electrodes positioned according to the international 10-20 system. Data were sampled at 5000 Hz and referenced online to Fp2, with the ground at AFz. We aimed to keep electrode impedances below 50 kΩ. The onset and offset of every trial were marked with a paired trigger.

#### Intervention Conditions

The two intervention protocols are those reported in Kubit et al. (2025), where they are called the gamma music-based intervention (gMBI) and the health education intervention (HEI), and where we described the feasibility and tolerability of both using natural language analyses of participants’ daily logbooks and of research assistants’ notes from weekly check-in calls. We refer to them here as the treatment and control protocols. The present report draws on the same logbooks for how long and how often participants listened, and adds the behavioral and electrophysiological outcomes.

##### Treatment Protocol: Music-Based Intervention

The treatment protocol paired visual stimulation with a self-administered, receptive, and individualized music listening program. We adapted that program from Hanser and Thompson (Hanser & Thompson, 1994), who reduced depression and anxiety in depressed older adults and found the benefit still present nine months later. The same program has since been used with older adult family caregivers and their relatives with dementia (Hanser et al., 2011) and with chronically ill patients (Roseen et al., 2017). As in the original, our version directs the participant’s attention to music they chose themselves, with the aim of strengthening positive emotional responses.

After enrolling, each participant met a member of the study team for a one-hour consultation and was compensated $20. During the meeting the participant and caregiver (if applicable) were shown how to use the light device and took it home for the eight weeks. The team member also asked what music the participant knew and liked, and sorted those pieces, along with music by similar artists or in similar genres and instrumentation, into two playlists: one intended to relax and one to energize. The playlists were loaded onto the participant’s own listening device. The participant chose a time of day to listen and a medium for it (earbuds, headphones or speakers).

Participants were asked to listen for at least 30 minutes a day across the eight weeks while viewing the light stimulation, choosing one of the two playlists each day. They were asked to attend closely to the music and to note anything they noticed in mind or body, including associations, memories, images, breathing rate, and tension or relaxation. After each session they recorded in a daily logbook how long they had listened, how relaxed and how energized the music made them feel, whether they enjoyed the lights, and any other effects. Logs were completed on paper and submitted either weekly in electronic form or as hard copy at the end of the intervention. A member of the study team called each participant weekly for approximately 30 minutes to check on use of the device and to choose playlists for the coming week.

##### Control Protocol: Health Education Intervention

The active control protocol matched the treatment protocol for duration, level of activity, and amount of contact with the study team. It differed in two respects: participants listened to health education podcasts rather than self-selected music, and the light stimulation was driven in the delta band only, without the theta and gamma components described under Materials. Control participants also met a team member for a one-hour consultation, were compensated $20 for it, were shown how to use the light device, and took it home for the eight weeks. In place of music playlists, they chose from a database of more than 100 health education podcasts selected for their age group, with a different topic each week. They listened for at least 30 minutes a day while viewing delta-driven light stimulation, and recorded notes about each podcast in the same daily logbook. A member of the study team called each participant weekly for approximately 30 minutes to check on compliance and to choose podcasts for the coming week.

#### Post-Intervention Assessment

Participants returned to the laboratory as soon as possible after completing the 8-week intervention. They repeated the full pre-intervention procedure and took part in an exit interview, in which they reflected on their time in the trial and gave feedback to the research team. The post-intervention session took approximately 7 hours, and participants were compensated $20 per hour. Responses from the exit interviews are not analyzed here.

### Materials

#### Light Stimulus

To deliver gamma-band stimulation visually rather than acoustically, we built a custom light device: a ring of LEDs positioned to subtend a 20° visual angle around the fovea. We chose that angle because retinotopic mapping work indicates it is well suited to detecting motion and small changes in brightness (Purves et al., 2013). The device took audio in, filtered the acoustic signal, and used it to drive a simulated network of neural oscillators that predicts the frequency, phase, and amplitude of human neural responses to that audio (Large et al., 2015). Those simulated oscillations drove the LEDs, pulsing on the beat and changing color on strong beats (Large et al., 2015; Tichko, Kim, et al., 2022). The delta frequencies the device extracts have been validated against annotated databases of musical examples (Hainsworth & Macleod, 2004; Janata et al., 2012).

In the treatment protocol the lights tracked the delta (1-4 Hz) and theta (4-8 Hz) components of the music, which correspond to the beat and its subdivision in most music (Ding et al., 2017; Doelling & Poeppel, 2015; Large et al., 2015). The same signal was additionally amplitude-modulated at a fixed 39 Hz, which produced a visible flicker above the beat-level modulation. We held the gamma rate fixed rather than deriving it from the music, so that every participant received stimulation at the same gamma frequency regardless of what they chose to listen to. In the control protocol the lights tracked delta only, with no theta component and no gamma modulation, so the beat-level modulation was visible and the flicker was absent. Both protocols used the same device. Crossmodal theta-gamma phase-amplitude coupling was present in our stimulus by design and, importantly, was missing from the control protocol’s delta-driven light stimulation.

### EEG Preprocessing

Preprocessing of EEG data used EEGLAB 2021.1 (Delorme & Makeig, 2004) in MATLAB 2024b. Electrode positions were assigned from the standard 10-05 template (Oostenveld & Praamstra, 2001). Data recorded before the first trial, between trials, and after the last trial were discarded, and the retained trial intervals were concatenated into a single continuous recording with a boundary marker at each splice. All subsequent preprocessing was applied to this single recording; the data were separated by condition only after artifact correction.

Data were downsampled to 500 Hz, high-pass filtered at 1 Hz, cleaned of 60 Hz line noise using CleanLine (Mullen, 2012; 2 Hz bandwidth, 4 s window, 1 s step, p < .01), then low-pass filtered at 55 Hz. Both filters were zero-phase, non-causal, Hamming-windowed sinc FIR filters applied with the default order estimate of pop_eegfiltnew (Widmann et al., 2015). Bad channels were identified with clean_rawdata. A channel was rejected if it correlated below .8 with a reconstruction of it from neighboring channels for more than 50% of the recording, or if its line noise exceeded 4 SD of its own signal. Artifact subspace reconstruction (ASR), which reconstructs noisy data epochs from a low-dimensional principal component subspace across all channels (Mullen et al., 2015), was used to correct high-variance bursts at a threshold of 10 SD; no time windows were removed. Rejected channels were then interpolated using spherical splines (Perrin et al., 1989). Data were re-referenced to the average of TP9 and TP10, leaving 30 channels for analysis. Across sessions, an average of 4 channels (range: 2-7) were rejected and interpolated.

ICA was computed on the continuous re-referenced data using the extended Infomax algorithm (runica; Bell & Sejnowski, 1995; Lee et al., 1999). Components were classified with ICLabel (Pion-Tonachini et al., 2019), each component was assigned to its most probable class, and every component whose most probable class was not classified as “Brain” or “Other” was subtracted. An average of 12 components (range: 4-21) were removed per session. The remaining data were re-referenced to the average of all channels. Finally, cleaned continuous data were cut into one segment per trial using the onset and offset trigger pairs, giving four stimulus segments per session, one for each combination of song and light condition.

### Statistical Analyses

This is an interim analysis of an ongoing randomized feasibility trial. Two behavioral outcomes were specified in advance: a decrease in ADAS-Cog score and an increase in face-name association score, both indicating improvement. Theta-gamma PAC was specified as the primary EEG outcome. The associations between coupling and cognition are additional analyses, and we report them without correction across measures. Behavioral analyses were conducted in R version 4.5.1 (2025-06-13). EEG analyses were conducted in MATLAB 2024b using EEGLAB 2021.1. Alpha was set at .05 and all tests were two-tailed. We report effect sizes throughout. Confidence intervals for the cluster effect size and for correlations were obtained by bootstrap resampling of participants; all other 95% intervals are parametric.

#### Compliance

We compared the two arms over the 8-week period with two-sample t-tests on the number of days a participant logged any listening, the mean length of a listening session, the number of days a participant watched and listened for more than the required 30 minutes, total listening time, and daily ratings of how enjoyable and how stimulating the lights were. For each we report the difference in means with its 95% confidence interval and Cohen’s *d*. Compliance measures came from the daily logbooks which participants completed by hand after each session. Days they did not listen were recorded as 0 minutes.

Logbook data are incomplete for four participants who completed the intervention and remained in contact with research staff throughout. These four were evenly split between groups, two in the treatment group and two in the control group. One lost their logbook entirely and is entered here as zero minutes. Two kept their own records on separate paper after the first week, and one recorded podcast titles without listening times; in each case the records could not be reliably transcribed, so only the first week is represented. All four are included in the compliance comparisons. Research assistants transcribed and digitized the logs and removed personally identifying information before analysis. Analyses of the free-text responses in the logs and the notes from the weekly check-in calls are ongoing and will be reported separately.

#### Stimulus Validation

We also checked, in the pre-intervention EEG session, that the music and light stimulation drove the neural oscillations it was built to engage. We first asked whether delta and theta activity in the EEG was phase-locked to the same frequencies in the music, with each participant’s values averaged across both light conditions. We then compared the delta-theta-gamma-driven light stimulation to the delta-driven light stimulation on delta, theta, and gamma gain, and on theta phase-locking value, using a paired cluster-based permutation test across the 30 electrodes, with each participant’s values averaged across the two songs (n = 33; one participant had no delta-driven recording for one song and was excluded from these tests). All cluster-based permutation tests were implemented in FieldTrip (ft_freqstatistics), with neighbors defined by triangulation of the electrode positions, electrodes entering a cluster at p < .05, 2,000 permutations, and clusters considered significant at a corrected p < .05. For each significant cluster we averaged gain or phase-locking value across the electrodes within each participant and condition, and reported d_z_ on the difference with a bootstrapped 95% confidence interval.

#### Behavioral Assessments

We evaluated change on each behavioral measure with a session (pre-intervention, post-intervention) by intervention (treatment, control) 2x2 mixed ANOVA, with session as a within-subject factor and intervention as a between-subject factor. A separate model was fitted for the face-name association memory score and the ADAS-Cog score. We report partial eta squared for each effect. Post-hoc analyses included paired t-tests of pre-to-post change within each group and a two-sample t-test comparing change scores between groups, both assuming equal variances. Within-group effect sizes are *d*_z_, the mean change divided by the standard deviation of the change scores. The between-group effect size is Cohen’s *d*, the difference in mean change divided by the pooled standard deviation of the change scores.

#### Theta-Gamma Phase-Amplitude Coupling

Theta-gamma PAC was present in our stimulus by design because the light stimulus amplitude-modulated a fixed 39 Hz carrier locked to the delta and theta components of the music. We measured whether the brain reproduces such coupling, across modalities, since the theta phase is carried by the audio and the gamma amplitude by the light. For each participant we computed a stimulus-driven theta-gamma PAC change score at each electrode by subtracting the pre-intervention value from the post-intervention value, averaged across the four listening trials in a session (two songs, each under delta-theta-gamma-driven and delta-driven light stimulation). We tested whether these change scores differed between intervention groups with a cluster-based permutation test across the 30 electrodes, using an independent-samples t statistic at each electrode and permuting intervention labels across participants. Cluster statistics were the summed t within a cluster, FieldTrip’s default. For the significant cluster we averaged PAC across its electrodes within each participant and session, and report Cohen’s d for the group difference in change with a 95% confidence interval.

#### Association Between Coupling and Memory

We tested the association between change in stimulus-driven theta-gamma PAC and change in face-name score across both groups with a Pearson correlation, assessed by permutation: we randomly reassigned participants’ PAC change values relative to their memory change values across 2,000 permutations and compared the observed correlation against the resulting null distribution. The 95% confidence interval was obtained by bootstrap resampling of participants over 2,000 resamples.

#### Electroencephalography

##### Gain

We asked whether our music and synchronized light stimulus entrained endogenous brain oscillations in our participants. To do this, we computed electroencephalography (EEG) power over the delta-theta-gamma ranges and identified response frequencies. We then computed local gain at the power peaks corresponding to the targeted frequencies (targeted frequencies differed depending on the music stimulus). We computed the neural response for each song separately because the specific stimulus frequencies differed for each song. We performed a time-frequency decomposition on the EEG signal, using a Constant-Q transform (cqt.m in MATLAB). The Constant-Q transform was chosen to provide the same relative bandwidth (i.e., constant-q) at delta, theta, and gamma frequencies. The result was then averaged over time to provide a frequency domain result, squared to compute power (µV^2^), and transformed to decibels (10 log_10_(µV^2^)). Gain was then measured as the difference between decibel values using an acausal FIR filter (second derivative of a Gaussian) that subtracts the power of surrounding frequency bands from the power at the frequency of interest.

##### Phase-Locking

When neural entrainment is driven by rhythms in an external stimulus, we expect that specific frequencies in the acoustic and EEG signals will show a consistent phase-lag (Obleser & Kayser, 2019). We capture this stimulus-brain phase alignment using the cerebro-acoustic phase-locking value (0 ≤ PLV ≤1), which we computed for each electrode and each song over a range of frequencies (0.2 - 20.2 Hz; step size: .2 Hz). First, we filtered the raw audio using a gammatone filterbank that mimics logarithmic filtering of the human cochlea using 10 logarithmically spaced filters (Lartillot et al., 2008). The cochlear-filtered acoustic signal was defined as the sum of the amplitude envelopes across filters. Second, we transformed the acoustic signal and EEG signal at each electrode into a complex time-frequency representation by convolving both signals with complex Morlet wavelets (5 cycles) defined for each center frequency in the range 0.2 - 20.2 Hz with a step size of .2 Hz. Third, we calculated the phase-locking value at each frequency by computing the mean vector length of the complex vectors defined by the phase difference between the EEG and acoustic phase timeseries.

##### Theta-Gamma Phase-Amplitude Coupling

We measured stimulus-driven theta to EEG-gamma phase-amplitude coupling (referred to as theta-gamma PAC) using the circular-linear correlation method from Reinhart & Nguyen (2019). We extracted the phase time series at the hypothesized theta frequency from the cochlear-filtered envelope of each song (4.09 Hz for “Under the Boardwalk”; 3.92 Hz for “Graceland”) and an amplitude time series at the hypothesized gamma frequency (39 Hz) for each electrode using complex Morlet wavelet decomposition. We defined the number of wavelet cycles as a linear function of frequency using the equation (s=freq/5+3), to balance the tradeoff between time- and frequency-domain resolution. This resulted in a wavelet full-width half-maximum (FWHM) in the frequency domain of ∼2.8 Hz for the theta frequency and ∼9.6 Hz for the gamma frequency. Importantly, the frequency tapering at the gamma frequency included the amplitude modulation sidebands of the theta frequency (FWHM(γ) greater than the theta frequency in either song), which is necessary for detecting phase-amplitude coupling (Aru et al., 2015). We obtained the final theta-gamma coupling value by computing the circular-linear correlation value between the theta phase series and the gamma amplitude series (Berens & Velasco, 2009) and performing Fisher’s z transformation on the resulting correlation coefficient.

## Data Availability

All data produced in the present study are available upon reasonable request to the authors.

## Acknowledgements

This work was supported by: NIH R01AG078376, NIH R21AG075232, NIH R43AG092067, NSF-BCS 2240330 and NSF-CAREER 1945436. We thank former lab members Parker Tichko, Milena Quinci, Sherry Wang, Nick Kathios, Yaen Chen, Xiyu Zhang, former research assistants Hannah Weibley, Quincy Dillard, Julia Rivas, Brad Hobert, and Serafina Kubit-Whiteside. We also thank recruitment site Newbridge on the Charles as well as Letitia Preto, Nancy Donovan, Amy Lo.

## Supplemental Methods

### Methods

#### Neuropsychological Assessments

Participants completed a battery of neuropsychological measures at both sessions, drawn from several instruments rather than from a single published battery. We describe each below in the order administered. A member of the study team administered all measures.

#### FAS Verbal Fluency Test

We measured verbal fluency with the FAS letter fluency test from the Neurosensory Center Comprehensive Examination for Aphasia (Spreen, 1977) together with a category fluency test. In the letter task the participant named as many words as possible beginning with F, then A, then S, with one minute for each letter, excluding proper nouns and repeated word stems. In the category task the participant named as many animals, then fruits, then vegetables as possible, again with one minute for each. The total score is the sum of correct words across all six trials, minus repetitions and set losses. Higher scores indicate better performance.

#### Trail Making Test

We measured visual scanning and set-shifting with the Trail Making Test (Armitage, 1946). In Part A the participant drew lines connecting 25 numbered circles in ascending order. In Part B the circles were numbered 1 to 13 and lettered A to L, and the participant alternated between the two sequences.

Both parts are scored as time to completion in seconds, so higher scores indicate worse performance. A completion time of over 273 seconds is indicative of a deficit.

#### Wechsler Memory Scale-Revised (WMS-R) Logical Memory

We measured verbal episodic memory with the Logical Memory subtest of the WMS-R (Wechsler, 1987). The examiner read two short stories aloud, the first once and the second twice, and the participant freely recalled each story immediately after hearing it. After approximately 30 minutes the participant recalled both stories again. Scores are the number of story units recalled, immediately (Logical Memory I) and after the delay (Logical Memory II). Higher scores indicate better performance.

#### Wechsler Adult Intelligence Scale - Fourth Edition (WAIS-IV) Coding Test

We measured processing speed with the Coding subtest of the WAIS-IV (Wechsler, 2008). The participant was shown a key pairing the digits 1 through 9 with symbols, and copied the matching symbol beneath as many digits as possible in 120 seconds. The score is the number of correctly copied symbols. Higher scores indicate better performance.

#### Wechsler Adult Intelligence Scale - Fourth Edition (WAIS-IV) Digit Span

We measured attention and working memory with the Digit Span subtest of the WAIS-IV (Wechsler, 2008). The examiner read sequences of digits aloud, which the participant repeated in the same order, in reverse order, and in ascending numerical order. The score is the total across all three conditions. Higher scores indicate better performance.

#### National Adult Reading Test (NART)

We measured premorbid verbal ability with the National Adult Reading Test (NART; (Nelson, 1982)). The participant read aloud a list of 50 irregularly spelled words, and was not told the correct pronunciation or whether a response was right or wrong. The score is the number pronounced correctly. Higher scores indicate better performance.

#### Hopkins Verbal Learning Test - Revised (HVLT-R)

We measured verbal learning and retention with the HVLT-R (Benedict et al., 1998). The examiner read a list of 12 words drawn from three semantic categories, and the participant freely recalled as many as possible; this was repeated across three learning trials. After a delay of 20 to 25 minutes the participant recalled the list again, and then completed a yes/no recognition trial of 24 words, 12 from the list and 12 foils. We used two scores: retention, the proportion of words recalled after the delay relative to the best of the second and third learning trials, and the recognition discrimination index, true positives minus false positives. Higher scores indicate better performance.

#### Scoring Reliability

Logical Memory, NART, and verbal fluency were scored independently by three members of the study team, and we computed intraclass correlations between scorers for each measure.

## Supplemental Results

### Compliance

**Figure S1.**
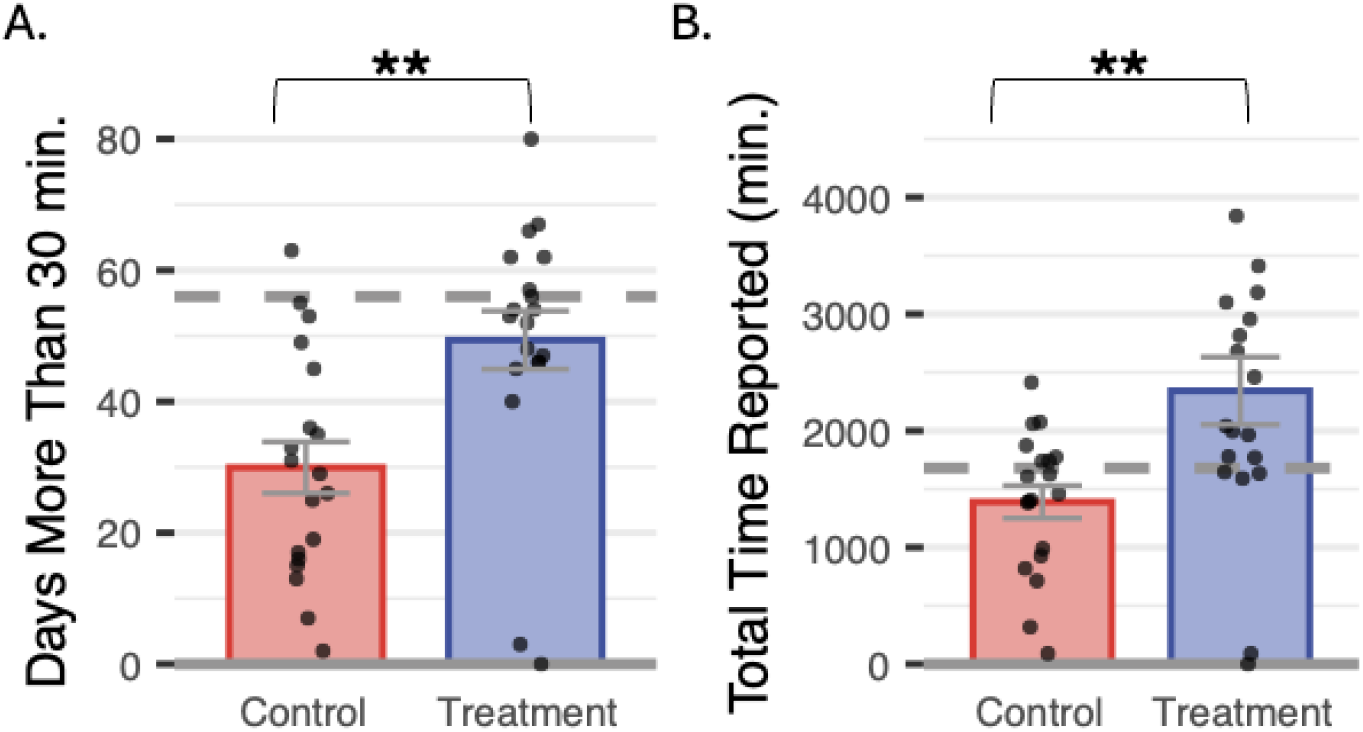
(A) Number of days in the intervention period that participants listened and watched for more than 30 minutes stratified by intervention group and (B) total listening and watching time throughout the entire 8-week intervention by intervention group. Asterisks indicate significance of t-tests (* *p* < .05; ** *p* < .01; *** *p* < .001).

### Behavioral

Table S1 gives pre- and post-intervention means and standard deviations for each neuropsychological measure by intervention group. Analyses of these measures are ongoing and will be reported separately; ten measures tested at two sessions in two groups would require a correction that the present interim sample cannot support.

**Table S1.** Neuropsychological measures before and after the 8-week intervention, by group.

|  | **Control** |  | **Treatment** |  |
|---|---|---|---|---|
| **Measure** | **Pre** | **Post** | **Pre** | **Post** |
| National Adult Reading Test | 32.1 (10.4) | 32.3 (9.4) | 27.7 (9.6) | 29.6 (9.8) |
| WMS-R Logical Memory I (immediate) | 37.4 (12.2) | 39.3 (10.7) | 31.6 (12.1) | 31.4 (11.9) |
| WMS-R Logical Memory II (delayed) | 21.3 (9.4) | 23.6 (9.0) | 17.1 (8.5) | 18.1 (8.3) |
| HVLT-R retention | 61.8 (26.8) | 67.5 (28.0) | 43.4 (34.0) | 59.2 (61.4) |
| HVLT-R recognition discrimination | 10.2 (1.9) | 10.1 (2.2) | 8.2 (3.0) | 8.2 (3.8) |
| WAIS-IV Digit Span | 27.7 (7.7) | 29.8 (9.3) | 24.7 (7.8) | 24.4 (7.5) |
| WAIS-IV Coding | 56.9 (16.8) | 59.4 (18.2) | 49.1 (14.3) | 50.3 (14.1) |
| Trail Making Test A | 37.7 (19.5) | 36.1 (24.2) | 38.1 (20.9) | 44.5 (44.5) |
| Verbal fluency (FAS and category) | 90.9 (25.6) | 94.2 (26.8) | 74.7 (25.5) | 75.0 (27.3) |
| Trail Making Test B | 92.3 (71.2) | 91.7 (80.7) | 128.8 (91.8) | 106.1 (89.2) |

**Note**. Values are means with standard deviations in parentheses. n = 38 (19 per group) except for the National Adult Reading Test, the HVLT-R recognition discrimination index, and WAIS-IV Digit Span, for which n = 37. Higher scores indicate better performance on all measures except the Trail Making Test, Parts A and B, which are scored as time to completion in seconds.

### EEG

The primary EEG analysis showed that the change in stimulus-driven theta to EEG-gamma phase-amplitude coupling over the course of the intervention was significantly stronger for the treatment group compared to the control group. The effect we observed on PAC aligns with a set of significant Intervention x Session interaction effects on EEG gain and phase-locking as well. Change from pre- to post-intervention differed between groups for delta gain (C3, CP5, Pz, P3, O1, Oz; cluster-corrected p = .011, within-cluster d = 1.31, 95% CI [0.68, 2.20]), theta gain (P3, O1, Oz, O2; cluster-corrected p = .013, within-cluster d = 1.60, 95% CI [1.02, 2.41]), and theta phase-locking (P7, O1, Oz, O2; cluster-corrected p = .029, within-cluster d = 0.96, 95% CI [0.34, 1.72]), in overlapping central-parietal and occipital clusters. In each case the measure increased in the treatment group and decreased in the control group.

Gamma gain did not significantly differ between groups (p = .285). These results suggest that the music listening intervention not only affected cross-frequency coupling, but also directly strengthened stimulus-driven delta and theta brain dynamics.

**Figure S2.**
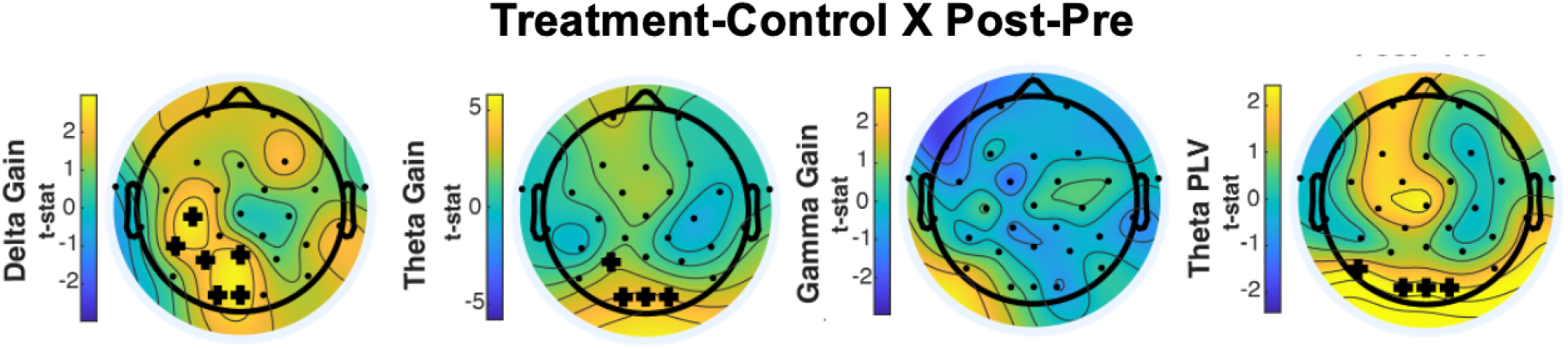
Topographic plots showing the interaction between session (pre- to post-intervention) and group (treatment versus control) on EEG gain and phase-locking at stimulated frequencies. Electrodes that showed a significant effect (p<.05; cluster-based permutation tests) are marked with “+”.

## References

1. 2025 Alzheimer’s disease facts and figures. (2025). Alzheimer’s & Dementia, 21(4), e70235. 10.1002/alz.70235

2. Adaikkan, C., Middleton, S. J., Marco, A., Pao, P.-C., Mathys, H., Kim, D. N.-W., Gao, F., Young, J. Z., Suk, H.-J., Boyden, E. S., McHugh, T. J., & Tsai, L.-H. (2019). Gamma Entrainment Binds Higher-Order Brain Regions and Offers Neuroprotection. Neuron, 102(5), 929–943.e8. 10.1016/j.neuron.2019.04.011

3. Alekseichuk, I., Turi, Z., Amador de Lara, G., Antal, A., & Paulus, W. (2016). Spatial Working Memory in Humans Depends on Theta and High Gamma Synchronization in the Prefrontal Cortex. Current Biology, 26(12), 1513–1521. 10.1016/j.cub.2016.04.035

4. Armitage, S. G. (1946). Trail Making Test. 10.1037/t00757-000

5. Arnal, L. H., Kleinschmidt, A., Spinelli, L., Giraud, A.-L., & Mégevand, P. (2019). The rough sound of salience enhances aversion through neural synchronisation. Nature Communications, 10(1), 3671. 10.1038/s41467-019-11626-7

6. Aru, J., Aru, J., Priesemann, V., Wibral, M., Lana, L., Pipa, G., Singer, W., & Vicente, R. (2015). Untangling cross-frequency coupling in neuroscience. *Current Opinion in Neurobiology*, SI: Brain Rhythms and Dynamic Coordination, 31, 51–61. 10.1016/j.conb.2014.08.002

7. Bell, A. J., & Sejnowski, T. J. (1995). An information-maximization approach to blind separation and blind deconvolution. Neural Computation, 7(6), 1129–1159. 10.1162/neco.1995.7.6.1129

8. Benedict, R. H. B., Schretlen, D., Groninger, L., & Brandt, J. (1998). Hopkins Verbal Learning Test – Revised: Normative Data and Analysis of Inter-Form and Test-Retest Reliability. The Clinical Neuropsychologist, 12(1), 43–55. 10.1076/clin.12.1.43.1726

9. Berens, P., & Velasco, M. J. (2009). *The circular statistics toolbox for Matlab*. https://pure.mpg.de/rest/items/item_1789418/component/file_3009634/content

10. Bier, N., Van Der Linden, M., Gagnon, L., Desrosiers, J., Adam, S., Louveaux, S., & Saint-Mleux, J. (2008). Face–name association learning in early Alzheimer’s disease: A comparison of learning methods and their underlying mechanisms. Neuropsychological Rehabilitation, 18(3), 343–371. 10.1080/09602010701694723

11. Bolland, E., De Burca, A., Wang, S. H., Khalil, A., & McLoughlin, G. (2025). Efficacy of auditory gamma stimulation for cognitive decline: A systematic review of individual and group differences across cognitively impaired and healthy populations. Npj Aging, 12(1), 8. 10.1038/s41514-025-00305-1

12. Brandt, J., Spencer, M., & Folstein, M. (1988). The telephone interview for cognitive status. Neuropsychiatry Neuropsychol Behav Neurol, 1(2), 111–117.

13. Canolty, R. T., & Knight, R. T. (2010). The functional role of cross-frequency coupling. Trends in Cognitive Sciences, 14(11), 506–515. 10.1016/j.tics.2010.09.001

14. Cardona, G., Ferreri, L., Lorenzo-Seva, U., Russo, F. A., & Rodriguez-Fornells, A. (2022). The forgotten role of absorption in music reward. Annals of the New York Academy of Sciences, 1514(1), 142–154. 10.1111/nyas.14790

15. Chan, D., Suk, H.-J., Jackson, B. L., Milman, N. P., Stark, D., Klerman, E. B., Kitchener, E., Avalos, V. S. F., Weck, G. de, Banerjee, A., Beach, S. D., Blanchard, J., Stearns, C., Boes, A. D., Uitermarkt, B., Gander, P., Iii, M. H., Sternberg, E. J., Nieto-Castanon, A., … Tsai, L.-H. (2022). Gamma frequency sensory stimulation in mild probable Alzheimer’s dementia patients: Results of feasibility and pilot studies. PLOS ONE, 17(12), e0278412. 10.1371/journal.pone.0278412

16. Chen, W. G., Iversen, J. R., Kao, M. H., Loui, P., Patel, A. D., Zatorre, R. J., & Edwards, E. (2022). Music and Brain Circuitry: Strategies for Strengthening Evidence-Based Research for Music-Based Interventions. Journal of Neuroscience, 42(45), 8498–8507. 10.1523/JNEUROSCI.1135-22.2022

17. Clare, L., Wilson, B. A., Carter, G., Roth, I., & Hodges, J. R. (2002). Relearning face-name associations in early Alzheimer’s disease. Neuropsychology, 16(4), 538–547. 10.1037/0894-4105.16.4.538

18. Cui, T., Han, X., Yao, Q., Xu, C., Shan, A., Wang, X., Li, S., Luan, H., Wen, B., Chen, R., Guo, J., Lian, Q., Li, C., Sun, Y., Lv, S., & Wei, C. (2026). Clinical efficacy of non-pharmacological interventions on cognitive improvement in patients with mild to moderate Alzheimer’s disease: A systematic review and Bayesian network meta-analysis. Journal of Alzheimer’s Disease, 13872877261485496. 10.1177/13872877261485496

19. Delorme, A., & Makeig, S. (2004). EEGLAB: An open source toolbox for analysis of single-trial EEG dynamics including independent component analysis. Journal of Neuroscience Methods, 134(1), 9–21. 10.1016/j.jneumeth.2003.10.009

20. Ding, N., Patel, A. D., Chen, L., Butler, H., Luo, C., & Poeppel, D. (2017). Temporal modulations in speech and music. *Neuroscience & Biobehavioral Reviews*, The Biology of Language, 81, 181–187. 10.1016/j.neubiorev.2017.02.011

21. Doelling, K. B., & Poeppel, D. (2015). Cortical entrainment to music and its modulation by expertise. Proceedings of the National Academy of Sciences, 112(45), E6233–E6242. 10.1073/pnas.1508431112

22. Folstein, M. F., Folstein, S. E., & McHugh, P. R. (1975). Mini-Mental State Examination. 10.1037/t07757-000

23. Fujioka, T., Trainor, L. J., Large, E. W., & Ross, B. (2009). Beta and Gamma Rhythms in Human Auditory Cortex during Musical Beat Processing. Annals of the New York Academy of Sciences, 1169(1), 89–92. 10.1111/j.1749-6632.2009.04779.x

24. Goodman, M. S., Kumar, S., Zomorrodi, R., Ghazala, Z., Cheam, A. S. M., Barr, M. S., Daskalakis, Z. J., Blumberger, D. M., Fischer, C., Flint, A., Mah, L., Herrmann, N., Bowie, C. R., Mulsant, B. H., & Rajji, T. K. (2018). Theta-Gamma Coupling and Working Memory in Alzheimer’s Dementia and Mild Cognitive Impairment. Frontiers in Aging Neuroscience, 10. 10.3389/fnagi.2018.00101

25. Grover, S., Wen, W., Viswanathan, V., Gill, C. T., & Reinhart, R. M. G. (2022). Long-lasting, dissociable improvements in working memory and long-term memory in older adults with repetitive neuromodulation. Nature Neuroscience, 25(9), 1237–1246. 10.1038/s41593-022-01132-3

26. Hainsworth, S. W., & Macleod, M. D. (2004). Particle Filtering Applied to Musical Tempo Tracking. EURASIP Journal on Advances in Signal Processing, 2004(15), 927847. 10.1155/S1110865704408099

27. Hanser, S. B., Butterfield-Whitcomb, J., Kawata, M., & Collins, B. E. (2011). Home-based Music Strategies with Individuals who have Dementia and their Family Caregivers. Journal of Music Therapy, 48(1), 2–27. 10.1093/jmt/48.1.2

28. Hanser, S. B., & Thompson, L. W. (1994). Effects of a Music Therapy Strategy on Depressed Older Adults. Journal of Gerontology, 49(6), P265–P269. 10.1093/geronj/49.6.P265

29. Harding, E. E., Sammler, D., Henry, M. J., Large, E. W., & Kotz, S. A. (2019). Cortical tracking of rhythm in music and speech. NeuroImage, 185, 96–101. 10.1016/j.neuroimage.2018.10.037

30. Henry, M. J., Herrmann, B., & Obleser, J. (2014). Entrained neural oscillations in multiple frequency bands comodulate behavior. Proceedings of the National Academy of Sciences, 111(41), 14935–14940. 10.1073/pnas.1408741111

31. Heusser, A. C., Poeppel, D., Ezzyat, Y., & Davachi, L. (2016). Episodic sequence memory is supported by a theta–gamma phase code. Nature Neuroscience, 19(10), 1374–1380. 10.1038/nn.4374

32. Hillebrand, M. C., Weise, L., & Wilz, G. (2023). Immediate effects of individualized music listening on behavioral and psychological symptoms of dementia: A randomized controlled trial. International Journal of Geriatric Psychiatry, 38(3), e5893. 10.1002/gps.5893

33. Iaccarino, H. F., Singer, A. C., Martorell, A. J., Rudenko, A., Gao, F., Gillingham, T. Z., Mathys, H., Seo, J., Kritskiy, O., Abdurrob, F., Adaikkan, C., Canter, R. G., Rueda, R., Brown, E. N., Boyden, E. S., & Tsai, L.-H. (2016). Gamma frequency entrainment attenuates amyloid load and modifies microglia. Nature, 540(7632), 230–235. 10.1038/nature20587

34. Janata, P., Tomic, S. T., & Haberman, J. M. (2012). Sensorimotor coupling in music and the psychology of the groove. Journal of Experimental Psychology: General, 141(1), 54–75. 10.1037/a0024208

35. Jeste, D. V., Palmer, B. W., Appelbaum, P. S., Golshan, S., Glorioso, D., Dunn, L. B., Kim, K., Meeks, T., & Kraemer, H. C. (2007). A new brief instrument for assessing decisional capacity for clinical research. Archives of General Psychiatry, 64(8), 966–974. 10.1001/archpsyc.64.8.966

36. Kubit, B. M., Parrish, C., Zhao, Z., & Loui, P. (2025). Diary Analysis of an RCT: Natural Language Analyses of Gamma-Music-Based Intervention. Music and Medicine, 17(1), 10–16. 10.47513/mmd.v17i1.1066

37. Large, E. W., Herrera, J. A., & Velasco, M. J. (2015). Neural Networks for Beat Perception in Musical Rhythm. Frontiers in Systems Neuroscience, 9. 10.3389/fnsys.2015.00159

38. Lartillot, O., Toiviainen, P., & Eerola, T. (2008). A Matlab Toolbox for Music Information Retrieval. In C. Preisach, H. Burkhardt, L. Schmidt-Thieme, & R. Decker (Eds.), Data Analysis, Machine Learning and Applications (pp. 261–268). Springer. 10.1007/978-3-540-78246-9_31

39. Lee, T. W., Girolami, M., & Sejnowski, T. J. (1999). Independent component analysis using an extended infomax algorithm for mixed subgaussian and supergaussian sources. Neural Computation, 11(2), 417–441. 10.1162/089976699300016719

40. Lisman, J. E., & Jensen, O. (2013). The Theta-Gamma Neural Code. Neuron, 77(6), 1002–1016. 10.1016/j.neuron.2013.03.007

41. Loui, P., Peelle, J., & Grahn, J. (2026). Music-Based Interventions for Cognition and Brain Health: The ARM framework. https://osf.io/preprints/psyarxiv/2mfzk

42. Martorell, A. J., Paulson, A. L., Suk, H.-J., Abdurrob, F., Drummond, G. T., Guan, W., Young, J. Z., Kim,

43. D. N.-W., Kritskiy, O., Barker, S. J., Mangena, V., Prince, S. M., Brown, E. N., Chung, K., Boyden, E. S., Singer, A. C., & Tsai, L.-H. (2019). Multi-sensory Gamma Stimulation Ameliorates Alzheimer’s-Associated Pathology and Improves Cognition. Cell, 177(2), 256–271.e22. 10.1016/j.cell.2019.02.014

44. Mas-Herrero, E., Marco-Pallares, J., Lorenzo-Seva, U., Zatorre, R. J., & Rodriguez-Fornells, A. (2013). Individual Differences in Music Reward Experiences. Music Perception, 31(2), 118–138. 10.1525/mp.2013.31.2.118

45. Mohs, R. C., Knopman, D., Petersen, R. C., Ferris, S. H., Ernesto, C., Grundman, M., Sano, M., Bieliauskas, L., Geldmacher, D., Clark, C., & Thal, L. J. (1997). Development of cognitive instruments for use in clinical trials of antidementia drugs: Additions to the Alzheimer’s disease assessment scale that broaden its scope. Alzheimer Disease and Associated Disorders, 11(SUPPL. 2), S13–S21.

46. Morris, J. C. (1997). Clinical Dementia Rating: A Reliable and Valid Diagnostic and Staging Measure for Dementia of the Alzheimer Type. International Psychogeriatrics, 9, 173–176. 10.1017/S1041610297004870

47. Mullen, T. (2012). CleanLine EEGLAB plugin. *San Diego*, CA: Neuroimaging Informatics Toolsand Resources Clearinghouse (NITRC).

48. Mullen, T. R., Kothe, C. A. E., Chi, Y. M., Ojeda, A., Kerth, T., Makeig, S., Jung, T.-P., & Cauwenberghs, G. (2015). Real-Time Neuroimaging and Cognitive Monitoring Using Wearable Dry EEG. IEEE Transactions on Bio-Medical Engineering, 62(11), 2553–2567. 10.1109/TBME.2015.2481482

49. Müllensiefen, D., Gingras, B., Musil, J., & Stewart, L. (2014). The Musicality of Non-Musicians: An Index for Assessing Musical Sophistication in the General Population. PLOS ONE, 9(2), e89642. 10.1371/journal.pone.0089642

50. Murdock, M. H., Yang, C.-Y., Sun, N., Pao, P.-C., Blanco-Duque, C., Kahn, M. C., Kim, T., Lavoie, N. S., Victor, M. B., Islam, M. R., Galiana, F., Leary, N., Wang, S., Bubnys, A., Ma, E., Akay, L. A., Sneve, M., Qian, Y., Lai, C., … Tsai, L.-H. (2024). Multisensory gamma stimulation promotes glymphatic clearance of amyloid. Nature, 627(8002), 149–156. 10.1038/s41586-024-07132-6

51. Nelson, H. (1982). National Adult Reading Test (NART): For the Assessment of Premorbid Intelligence in Patients with Dementia: Test Manual. Windsor: NFER-Nelson. https://cir.nii.ac.jp/crid/1370846644342763143

52. Obleser, J., & Kayser, C. (2019). Neural Entrainment and Attentional Selection in the Listening Brain. Trends in Cognitive Sciences, 23(11), 913–926. 10.1016/j.tics.2019.08.004

53. Oostenveld, R., & Praamstra, P. (2001). The five percent electrode system for high-resolution EEG and ERP measurements. Clinical Neurophysiology: Official Journal of the International Federation of Clinical Neurophysiology, 112(4), 713–719. 10.1016/s1388-2457(00)00527-7

54. Pereira, C. S., Teixeira, J., Figueiredo, P., Xavier, J., Castro, S. L., & Brattico, E. (2011). Music and Emotions in the Brain: Familiarity Matters. PLOS ONE, 6(11), e27241. 10.1371/journal.pone.0027241

55. Perrin, F., Pernier, J., Bertrand, O., & Echallier, J. F. (1989). Spherical splines for scalp potential and current density mapping. Electroencephalography and Clinical Neurophysiology, 72(2), 184–187. 10.1016/0013-4694(89)90180-6

56. Pion-Tonachini, L., Kreutz-Delgado, K., & Makeig, S. (2019). ICLabel: An automated electroencephalographic independent component classifier, dataset, and website. NeuroImage, 198, 181–197. 10.1016/j.neuroimage.2019.05.026

57. Polcher, A., Frommann, I., Koppara, A., Wolfsgruber, S., Jessen, F., & Wagner, M. (2017). Face-Name Associative Recognition Deficits in Subjective Cognitive Decline and Mild Cognitive Impairment. Journal of Alzheimer’s Disease, 56(3), 1185–1196. 10.3233/JAD-160637

58. Purves, D., Cabeza, R., Huettel, S. A., LaBar, K. S., Platt, M. L., & Woldorff, M. G. (2013). Principles of cognitive neuroscience (Second edition.). Sinauer Associates.

59. Quinci, M. A., Belden, A., Goutama, V., Gong, D., Hanser, S., Donovan, N. J., Geddes, M., & Loui, P. (2022). Longitudinal changes in auditory and reward systems following receptive music-based intervention in older adults. Scientific Reports, 12(1), 11517. 10.1038/s41598-022-15687-5

60. Reinhart, R. M. G., & Nguyen, J. A. (2019). Working memory revived in older adults by synchronizing rhythmic brain circuits. Nature Neuroscience, 22(5), 820–827. 10.1038/s41593-019-0371-x

61. Rentz, D. M., Amariglio, R. E., Becker, J. A., Frey, M., Olson, L. E., Frishe, K., Carmasin, J., Maye, J. E., Johnson, K. A., & Sperling, R. A. (2011). Face-name associative memory performance is related to amyloid burden in normal elderly. Neuropsychologia, 49(9), 2776–2783. 10.1016/j.neuropsychologia.2011.06.006

62. Roseen, E. J., Cornelio-Flores, O., Lemaster, C., Hernandez, M., Fong, C., Resnick, K., Wardle, J., Hanser, S., & Saper, R. (2017). Inpatient Massage Therapy Versus Music Therapy Versus Usual Care: A Mixed-methods Feasibility Randomized Controlled Trial. Global Advances in Health and Medicine, 6, 2164957X17735816. 10.1177/2164957X17735816

63. Rosen, W. G., Mohs, R. C., & Davis, K. L. (1984). A new rating scale for Alzheimer’s disease. The American Journal of Psychiatry, 141(11), 1356–1364. 10.1176/ajp.141.11.1356

64. Salimpoor, V. N., van den Bosch, I., Kovacevic, N., McIntosh, A. R., Dagher, A., & Zatorre, R. J. (2013). Interactions Between the Nucleus Accumbens and Auditory Cortices Predict Music Reward Value. Science, 340(6129), 216–219. 10.1126/science.1231059

65. Singer, A. C., Martorell, A. J., Douglas, J. M., Abdurrob, F., Attokaren, M. K., Tipton, J., Mathys, H., Adaikkan, C., & Tsai, L.-H. (2018). Noninvasive 40-Hz light flicker to recruit microglia and reduce amyloid beta load. Nature Protocols, 13(8), 1850–1868. 10.1038/s41596-018-0021-x

66. Sloboda, J. A., O’Neill, S. A., & Ivaldi, A. (2001). Functions of Music in Everyday Life: An Exploratory Study Using the Experience Sampling Method. Musicae Scientiae, 5(1), 9–32. 10.1177/102986490100500102

67. Snyder, J. S., & Large, E. W. (2005). Gamma-band activity reflects the metric structure of rhythmic tone sequences. Cognitive Brain Research, 24(1), 117–126. 10.1016/j.cogbrainres.2004.12.014

68. Soula, M., Martín-Ávila, A., Zhang, Y., Dhingra, A., Nitzan, N., Sadowski, M. J., Gan, W.-B., & Buzsáki, G. (2023). Forty-hertz light stimulation does not entrain native gamma oscillations in Alzheimer’s disease model mice. Nature Neuroscience, 26(4), 570–578. 10.1038/s41593-023-01270-2

69. Sperling, R. A., Bates, J. F., Cocchiarella, A. J., Schacter, D. L., Rosen, B. R., & Albert, M. S. (2001). Encoding novel face-name associations: A functional MRI study. Human Brain Mapping, 14(3), 129–139. 10.1002/hbm.1047

70. Spreen, O. (1977). Neurosensory center comprehensive examination for aphasia. Neuropsychological Laboratory. https://cir.nii.ac.jp/crid/1573387449448531328

71. Tenchov, R., Sasso, J. M., & Zhou, Q. A. (2024). Alzheimer’s Disease: Exploring the Landscape of Cognitive Decline. ACS Chemical Neuroscience, 15(21), 3800–3827. 10.1021/acschemneuro.4c00339

72. Tichko, P., Kim, J. C., Large, E., & Loui, P. (2022). Integrating music-based interventions with Gamma-frequency stimulation: Implications for healthy ageing. European Journal of Neuroscience, 55(11–12), 3303–3323. 10.1111/ejn.15059

73. Tichko, P., Page, N., Kim, J. C., Large, E. W., & Loui, P. (2022). Neural Entrainment to Musical Pulse in Naturalistic Music Is Preserved in Aging: Implications for Music-Based Interventions. Brain Sciences, 12(12), Article 12. 10.3390/brainsci12121676

74. Tort, A. B. L., Komorowski, R. W., Manns, J. R., Kopell, N. J., & Eichenbaum, H. (2009). Theta–gamma coupling increases during the learning of item–context associations. Proceedings of the National Academy of Sciences, 106(49), 20942–20947. 10.1073/pnas.0911331106

75. Traikapi, A., & Konstantinou, N. (2021). Gamma Oscillations in Alzheimer’s Disease and Their Potential Therapeutic Role. Frontiers in Systems Neuroscience, 15. 10.3389/fnsys.2021.782399

76. Van Der Steen, J. T., Van Der Wouden, J. C., Methley, A. M., Smaling, H. J. A., Vink, A. C., & Bruinsma, M. S. (2025). Music-based therapeutic interventions for people with dementia. Cochrane Database of Systematic Reviews, 2025(3). 10.1002/14651858.CD003477.pub5

77. Wechsler, D. (1987). Wechsler memory scale-revised. *Psychological Corporation*. https://cir.nii.ac.jp/crid/1573105974508393472

78. Wechsler, D. (2008). Wechsler Adult Intelligence Scale*—Fourth Edition*. 10.1037/t15169-000

79. Widmann, A., Schröger, E., & Maess, B. (2015). Digital filter design for electrophysiological data—A practical approach. Journal of Neuroscience Methods, 250, 34–46. 10.1016/j.jneumeth.2014.08.002

80. Zatorre, R. J. (2015). Musical pleasure and reward: Mechanisms and dysfunction. Annals of the New York Academy of Sciences, 1337(1), 202–211. 10.1111/nyas.12677

